# Viral-based individualized neoantigen vaccine as adjuvant treatment in resected head and neck squamous cell carcinoma: immunogenicity and efficacy from a randomized Phase I trial

**DOI:** 10.64898/2026.01.06.25342687

**Authors:** C. Ottensmeier, J-P. Delord, A. Lalanne, C. Jamet, A-L. Le Gac, K. Bidet-Huang, B. Grellier, J. Deforges, M. Brandely, E. Quéméneur, B. Bastien, A. Tavernaro, G. Lacoste, V. Schoettel, C. Spring-Giusti, N. Silvestre, J.B Marchand, S. Robin, E. Dochy, M. Ceppi, A. Riva, N. Yamagata, P. Brattas, K. Onoguchi, Y. Yamashita, H. Fontenelle, M. Eggert Martinez, O. Baker, T. Jones, A. Schache, E. Piaggio, K. Bendjama, O. Lantz, C. Le Tourneau

## Abstract

In approximately one third of patients, resected head and neck squamous cell carcinoma will recur. We postulated that the induction of tumor neoantigen-specific T cell responses could prevent relapse. To this end, we developed TG4050, an individualized neoantigen therapeutic vaccine encoding up to 30 patient-specific predicted tumor neoantigens delivered by a Modified Vaccinia Ankara virus viral vector. We tested TG4050 as single agent in a randomized phase I trial. We found that of 16 evaluable patients randomized to immediate vaccination with TG4050, none relapsed after a median follow-up of 30 months, while 3 relapsed in the 16 control arm patients randomized to observation and treatment with TG4050 after recurrence. Polyepitopic responses to vaccine neoantigens were detected in the blood of patients from both arms after treatment initiation. These responses were maintained throughout treatment and persisted for over one year after the last dose. Vaccine neoantigen-specific CD8^+^ T cells had an effector phenotype and displayed high expression of cytotoxic and tissue-resident markers. TCR repertoire analysis showed that vaccine neoantigen-specific CD8^+^ T cell responses were polyclonal and comprised both *de novo* responses and amplification of pre-existing tumor-infiltrating T cell clones. Together, this translational data is consistent with the model in which single-agent delivery of TG4050 induces long-lasting tumor neoantigen-specific cytotoxic T cell responses that prevent tumor recurrence.

## Introduction

Head and neck cancer (HNC) is the sixth most frequently occurring cancer worldwide. In 2022, HNC accounted for 891,000 new cases and 458,000 deaths (Bray et al, 2024). Approximately 90% of HNCs are squamous cell carcinoma (HNSCC), which arise from the epithelial lining of the oral cavity, pharynx and larynx (Psyrri et al, 2014). Approximately 60% of patients with HNSCC are diagnosed with locally advanced disease. For more than two decades, the first line standard of care for resectable patients has been cisplatin-based chemoradiotherapy for patients who do not receive surgery, or surgery followed by risk adapted radiotherapy with or without chemotherapy (Machiels et al, 2020, NCCN Clinical Practice Guidelines in Oncology, 2023). Key factors linked to unfavorable prognosis include advanced tumor stage, Human Papillomavirus (HPV) negative status, continued tobacco/alcohol exposure, TP53 mutations, EGFR pathway alterations and pathological features such as extracapsular spread of lymph nodes, positive margins, lymphovascular or perineural invasion, and multiple lymph nodes metastases (Haque et al, 2019, Basyuni et al, 2022, Nair et al, 2022).

HNSCC patients have limited sensitivity to immune checkpoints inhibitors (ICIs) as demonstrated by the 13.3% nivolumab overall response rate (ORR) to second line nivolumab treatment in the CHECKMATE 141 study (Ferris et al, 2016) or by the 17% ORR in the intent-to-treat patient’s population treated with pembrolizumab in the first-line KEYNOTE-048 study (Burtness et al, 2019). Recently, two Phase 3 clinical trials reported significant improvements in resectable/resected locally advanced HNSCC. KEYNOTE-689 (Uppaluri et al, 2025) showed that pembrolizumab as a peri-operative treatment regimen increased event-free survival (EFS) by 27% in the intent-to-treat population. The NivoPostOp study (Bourhis et al, 2025) reported a 24% improvement in disease-free survival (DFS) using nivolumab in the post-operative setting. Nonetheless, not all patients benefit from this approach and additional progress is needed to further enhance patient outcome.

HNSCC presents as a highly heterogenous malignancy exhibiting large variability in tumor microenvironment (TME) composition and immune infiltration. In HPV-negative HNSCC, an immune-deserted TME is present in approximately two thirds of patients, whereas one third of patients present with an immunosuppressive TME characterized by myeloid infiltrates (Johnson et al, 2023, Mito et al, 2021). This heterogeneity is an obstacle to conventional off-the-shelf immunotherapy. Nevertheless, the extent of pre-existing T cell responses in the tumor holds significant prognostic value (Torri et al, 2024). Recent reports from HSNCC patients treated with both CT and ICI demonstrated that the quality and clonal diversity of T cell responses could predict clinical outcome (Paschold et al, 2025, Wang et al, 2025, Lai et al, 2025, Zhao et al, 2025). Multiple datasets now demonstrate that neoantigens, derived from mutational damage in the tumor cells and presented to T cells on MHC molecules, can be highly immunogenic as they are not subject to central tolerance in the thymus (Xie et al, 2023). Together, this suggests that immunotherapies capable of inducing T-cell responses against the tumor-specific neoepitopes derived from the tumor mutanome may be a promising therapeutic avenue for HNSCC.

To this end, we developed TG4050, a Modified Vaccinia Ankara virus (MVA)-based individualized neoantigen therapeutic vaccine (INTV) aimed at eliciting patient tumor-specific T cell responses. TG4050 is designed for each patient by comparing somatic mutations in the tumor with matching healthy tissue. Up to 30 neoantigens, selected for their predicted expression and immunogenicity, are encoded within an MVA viral vector and administered to the patient. The use of an MVA vector system is expected to provide broad immune stimulation signals to support the generation of strong and long-lasting T cell responses to the encoded neoantigens (Bendjama et al, 2017).

We hypothesized that TG4050 could trigger an adaptive immune response against tumor antigens and prevent relapses in patients with high risk locally advanced resected HNSCC. We evaluated TG4050 as adjuvant monotherapy in a randomized Phase I clinical trial in resected HNSCC patients with a complete response after surgery followed by (chemo-)radiation (NCT04183166). The primary objective of this trial was to assess TG4050 safety and tolerability. The secondary objectives were feasibility and clinical efficacy assessment by measuring disease free survival (DFS). Translational objectives included characterization of the neoantigen-specific T cell responses to treatment.

## Results

### Patients, study design and feasibility

The study included patients with newly diagnosed, locally advanced, HPV-negative, resectable HNSCC. Patients underwent surgery followed by adjuvant radiotherapy (RT) with or without cisplatin treatment. Patients were randomized upon confirmation of complete response by imaging, 3 months after completion of adjuvant treatment. A total of 80 participants were screened, of which 47 patients were screen failure (Fig 1a). Reasons for exclusion included: not undergoing surgery (total 4 patients including 2 for withdrawal of consent, 9%), early relapse after surgery (15 patients, 32%), withdrawal of consent or loss to follow up (9 and 3 patients respectively, 26% combined), ineligibility based on tumor characteristics (5 patients with pathological stage I/II, 2 HPV positive, 1 patient with no adequate tumor material for sequencing, 17% combined), decision to not administer adjuvant therapy (4 patients, 9%), intercurrent illness (1 patient, 2%), and failure to produce the investigational medicinal product (3 patients, 6%).

**Figure 1.**
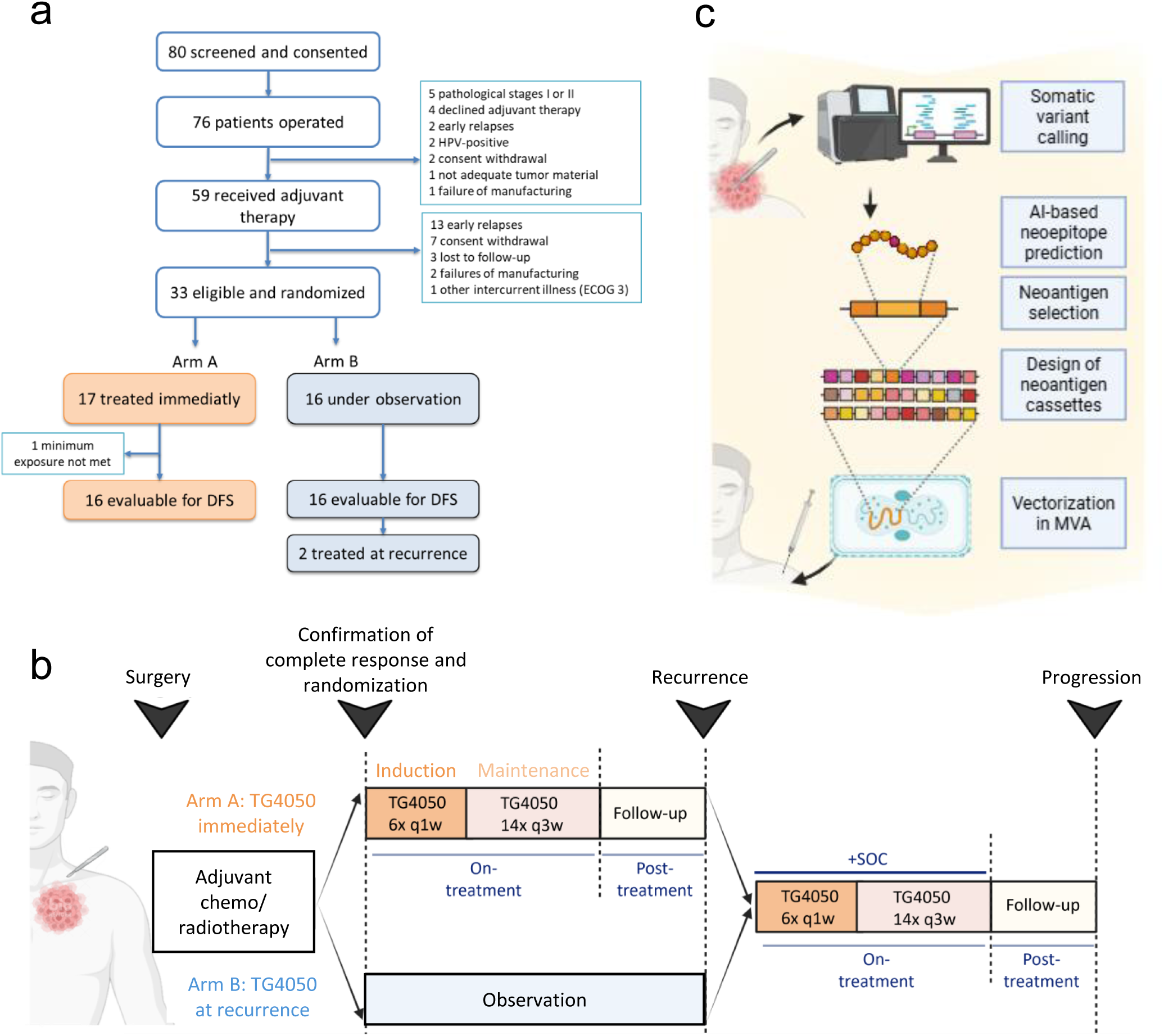
Study overview. **a**, CONSORT diagram. DFS: disease-free survival **b**, Phase 1 study design. **c**, TG4050 vaccine design: up to 30 neoantigens containing MHC class I and/or class II neoepitopes predicted from patient-specific somatic mutations are arranged in 3 optimized neoantigen cassettes and vectorized in Modified Vaccinia Ankara virus (MVA). Created with BioRender.

Thirty-three (33) participants were randomized between January 2021 and April 2023 across 3 sites in France and the United Kingdom. Patients were randomized to Arm A or B and treated according to the schedule in Fig 1b. Patients in Arm A received TG4050 within one week of randomization (thereafter referred as treated immediately), with 20 subcutaneous injections of TG4050 starting with an induction phase of 6 weekly doses followed by a maintenance phase of 14 doses at 3-week intervals. Patients in Arm B were kept under observation. In both arms, the same series of TG4050 could be given in combination with standard of care (SOC) at relapse.

Seventeen (17) patients were allocated to Arm A and 16 patients were allocated to Arm B (Intent-to-Treat population). One patient in Arm A was excluded from the Per Protocol population following the predefined criteria (Supplementary Table 1, minimum exposure not met). TG4050 was administered in combination with SOC after relapse in 2 patients in Arm B and in the early relapsing patient in Arm A.

Baseline demographic and disease characteristics were balanced between the treatment arms (Table 1). The majority of patients had a primary tumor location in the oral cavity and a pathological stage IVA or IVB. We collected information about positive resection margins, extracapsular extension, invaded lymph nodes, or perineural infiltration as key pathological risk factors. Patients in Arm A predominantly presented with 2 or more risk factors (9/17 vs 5/16 in Arm B), whereas more patients in Arm B presented with one risk factor (9/16 vs 5/17 in Arm A). More patients received chemoradiation than radiotherapy alone as adjuvant treatment. Tumor profiling based on next generation sequencing (NGS) analysis (Tumor Portrait^TM^, Supplementary Table 2) revealed a median tumor mutational burden (TMB) of 3.02 Mut/Mb, with a range of 0.03 –8.0 Mut/Mb (Table 1). Primary tumors were categorized according to the abundance of immune cells and fibroblasts according to the criteria described in (Bageav et al, 2021). The immune deserted TME was most common (20 patients, 61%), followed by non-fibrotic immune enriched (8 patients, 24%), fibrotic (3 patients, 9%) and fibrotic immune-enriched (2 patients, 6%).

**Table 1.** Baseline characteristics in the intent-to-treat (ITT) and per-protocol populations. Per protocol population: Minimum exposure before recurrence in Arm A: patients who completed their 6 weekly injections of TG4050, in Arm B: patients who completed their 6 weeks follow-up

| Characteristics | ITT population |  | Per-Protocol population |  |
| --- | --- | --- | --- | --- |
|  | Arm A<br>TG4050<br>immediately<br>(N=17) | Arm B<br>TG4050 at relapse<br>(N=16) | Arm A<br>TG4050 immediately<br>(N=16) | Arm B<br>TG4050 at relapse<br>(N=16) |
| Median age (range)-years | 61.0 (26-78) | 56.0 (47-73) | 61.5 (26-78) | 56.0 (47-73) |
| Male-no. (%) | 11 (64.7) | 13 (81.3) | 10 (62.5) | 13 (81.3) |
| ECOG PS 0-no. (%) | 12 (70.6) | 9 (56.3) | 12 (75.0) | 9 (56.3) |
| Stage at Diagnosis-no. (%)* |  |  |  |  |
| III | 7 (41.2) | 3 (18.8) | 7 (43.8) | 3 (18.8) |
| IVA | 10 (58.8) | 13 (81.3) | 9 (56.3) | 13 (81.3) |
| Pathological Stage-no. (%)* |  |  |  |  |
| I/II | 1 (5.9) | 1 (6.3) | 1 (6.3) | 1 (6.3) |
| III | 4 (23.5) | 3 (18.8) | 4 (25.0) | 3 (18.8) |
| IVA | 7 (41.2) | 5 (31.3) | 6 (37.5) | 5 (31.3) |
| IVB | 5 (29.4) | 7 (43.8) | 5 (31.3) | 7 (43.8) |
| Primary tumor site-no. (%) |  |  |  |  |
| Oral Cavity | 14 (82.4) | 10 (62.5) | 14 (87.5) | 10 (62.5) |
| Hypopharynx | 1 (5.9) | 3 (18.8) | 1 (6.3) | 3 (18.8) |
| Oropharynx | 2 (11.8) | 2 (12.5) | 1 (6.3) | 2 (12.5) |
| Larynx | 0 (0.0) | 1 (6.3) | 0 (0.0) | 1 (6.3) |
| Adjuvant treatment-no. (%) | 17 (100.0) | 16 (100.0) | 16 (100.0) | 16 (100.0) |
| Chemoradiation | 10 (58.8) | 9 (56.3) | 9 (56.3) | 9 (56.3) |
| Risk factors-no. (%) |  |  |  |  |
| Extracapsular extension | 5 (29.4) | 8 (50.0) | 5 (31.3) | 8 (50.0) |
| Positive resection margin | 4 (23.5) | 3 (18.8) | 4 (25.0) | 3 (18.8) |
| Perineural infiltration | 13 (76.5) | 7 (43.8) | 12 (75.0) | 7 (43.8) |
| Invaded lymph nodes ≥ 4 | 2 (11.8) | 3 (18.8) | 1 (6.3) | 3 (18.8) |
| Number of risk factors-no. (%) |  |  |  |  |
| 0 | 3 (17.6) | 2 (12.5) | 3 (18.8) | 2 (12.5) |
| 1 | 5 (29.4) | 9 (56.3) | 5 (31.3) | 9 (56.3) |
| 2 | 8 (47.1) | 3 (18.8) | 7 (43.8) | 3 (18.8) |
| 3 | 1 (5.9) | 2 (12.5) | 1 (6.3) | 2 (12.5) |
| Median TMB (range)-mut/Mb | 3.1 (1.4-7.7) | 3.0 (0.03-8.0) | 3.1 (1.4-7.7) | 3.0 (0.03-8.0) |
| Current or former smoker-no. (%) | 6 (35.3) | 6 (37.5) | 6 (37.5) | 6 (37.5) |
| Alcohol use, yes-no. (%) | 8 (47.1) | 7 (43.8) | 8 (50.0) | 7 (43.8) |

\*As defined by the American Joint Committee on Cancer eighth edition

TG4050 was designed for each patient based on tumor samples collected at surgery. First, tumor-specific somatic mutations were identified by comparing next generation DNA sequencing data from the resected tumor tissue with matched normal DNA obtained from blood as healthy tissue. Expression of the mutated antigens in the tumor tissue was confirmed by RNA sequencing. Neoepitopes were predicted using AI-based algorithms and up to 30 vaccine neoantigens (29-mers containing class I and/or class II epitopes) were selected for each patient based on expression, presentation and likelihood of T-cell recognition (Anzar et al, 2023). Expression cassettes were designed by arranging the neoantigens into 3 open reading frames (ORF) of up to 10 neoantigens each, using a proprietary algorithm to optimize manufacturability (Grellier et al, 2025). The resulting synthetic gene was cloned into an MVA viral vector and these recombinant vectors were produced under GMP conditions (Fig 1c).

TG4050 production was initiated for 76 patients after completion of surgery. Sequencing data could be obtained for all eligible patients but one, due to inadequate tumor material. Manufacturing was initiated for all patients with sequencing data and was stopped if patients were subsequently excluded from the study. Overall, TG4050 vaccines were successfully manufactured for 53 patients with a median number of 30 neoantigens (range 10-30). We were not able to manufacture a vaccine for 3 eligible patients.

### Safety and tolerability

Safety according to CTCAE v5 was analyzed in the Safety Population of 19 patients who received at least one injection of TG4050: 17 patients from Arm A and 2 patients receiving TG4050 after relapse in Arm B (**Table 2**). No dose-limiting toxicity was observed. No patients suffered grade 3 or more adverse events (AEs) related to TG4050. All patients but one (18/19, 94.7%) experienced grade 1/2 TG4050-related AEs which were, in most cases, injection site reactions (inflammation, pain, erythema, bruising, oedema, induration).

**Table 2.** Treatment-related Adverse Events (safety population)

|  | Arm A – TG4050 given before relapse<br>(N=17) |  |  | TG4050 given after relapse<sup>1</sup><br>(N=3) |  |  |
| --- | --- | --- | --- | --- | --- | --- |
|  | Grade 1<br>N (%) | Grade 2<br>N (%) | Grade 3+<br>N (%) | Grade 1<br>N (%) | Grade 2<br>N (%) | Grade 3+<br>N (%) |
| Treatment-related AEs |  |  |  |  |  |  |
| Patient(s) with at least one AE | 16 (94.1%) | 4 (23.5%) | 0 | 2 (66.7%) | 0 | 0 |
| Blood and lymphatic system disorders | 0 | 0 | 0 | 1 (33.3%) | 0 | 0 |
| Lymphopenia | 0 | 0 | 0 | 1 (33.3%) | 0 | 0 |
| Gastrointestinal disorders | 2 (11.8%) | 0 | 0 | 0 (0.0%) | 0 | 0 |
| Diarrhoea | 2 (11.8%) | 0 | 0 | 0 (0.0%) | 0 | 0 |
| General disorders and administration site conditions | 16 (94.1%) | 4 (23.5%) | 0 | 1 (33.3%) | 0 | 0 |
| Injections site reaction<sup>2</sup> | 16 (94.1%) | 3 (17.6%) | 0 | 1 (33.3%) | 0 | 0 |
| Fatigue | 2 (11.8%) | 0 | 0 | 0 (0.0%) | 0 | 0 |
| Influenza like illness | 1 (5.9%) | 1 (5.9%) | 0 | 0 (0.0%) | 0 | 0 |
| Investigations | 0 | 0 | 0 | 1 (33.3%) | 0 | 0 |
| Blood alkaline phosphatase increased | 0 | 0 | 0 | 1 (33.3%) | 0 | 0 |
| Nervous system disorders | 1 (5.9%) | 0 | 0 | 0 | 0 | 0 |
| Headache | 1 (5.9%) | 0 | 0 | 0 | 0 | 0 |
| Skin and subcutaneous tissue disorders | 2 (11.8%) | 1 (5.9%) | 0 | 0 | 0 | 0 |
| Rash | 2 (11.8%) | 1 (5.9%) | 0 | 0 | 0 | 0 |

<sup>1</sup> 1 patient in arm A and 2 patients in arm B received TG4050 in combination with SOC
<sup>2</sup> inflammation, pain, erythema, bruising, oedema, induration, pruritus, discolouration, swelling

### Clinical outcomes

Cut-off for efficacy data was 22 April 2025. Efficacy analyses were performed on the Per Protocol population, including 16 patients in each study arm. One patient in Arm A was excluded from the Per Protocol population due to early relapse after 2 doses of TG4050, which did not meet the minimum exposure defined per protocol. None of the 16 Arm A patients treated immediately after randomization have relapsed after a median follow-up of 30 months. Three of the 16 patients in Arm B relapsed while under observation: one patient after 6 months, one after 7 months and one after 18 months. The Kaplan – Meier estimates for DFS (Fig 2) were compared with a log-rank test (one-sided p value= 0.0367). The DFS rate at 2 years is 100% in Arm A and 81% in Arm B.

**Figure 2.**
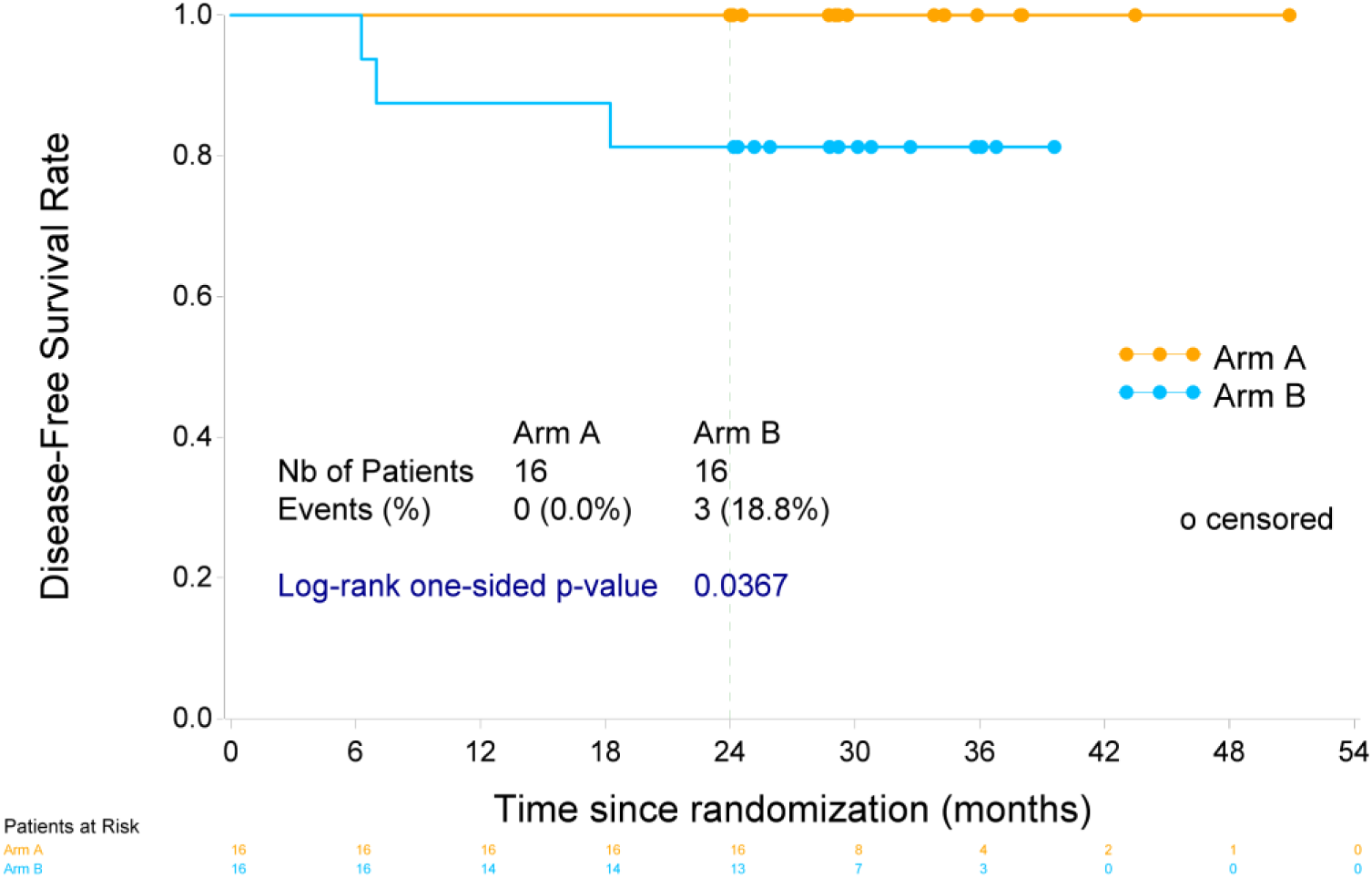
Kaplan-Meier estimates of disease-free survival in the per protocol population.

### Induction of neoantigen-specific T cell responses

In patients treated with TG4050 immediately, blood samples were collected by apheresis at baseline (before TG4050 first administration) and day 64 (D64, after 7 doses of TG4050) to assess the induction of circulating neoantigen-specific T cell responses. Longitudinal PBMC samples were obtained in the treatment induction and maintenance phase and up to 24 months (M24, one year after the end of treatment) to follow the dynamics of the response.

We used two complementary methods to detect T cell responses to treatment. To provide a functional breadth and magnitude readout for neoantigens inducing total (CD4^+^ and/or CD8^+^) T cell responses, we performed *ex vivo* IFN-γ ELISpot, systematically testing the response to each vaccine neoantigen. In parallel, we examined neoepitope-specific CD8^+^ T cells using a selection of predicted peptide-MHC class I tetramers (hereafter referred as tetramers) to enable a sensitive readout of the minimal epitope recognized and phenotype of the CD8^+^ T cell response at the neoepitope level. For both methods, responses to treatment were defined as *de novo* if they were below detection level at baseline and above detection level at D64, or as amplified if they were already detected at baseline but expanded at D64.

*Ex vivo* IFN-γ ELISpot was performed at baseline and D64 for 13 Arm A patients with sufficient PBMC samples. T cell responses to at least one vaccine neoantigen were detected in 8/13 (61.5%) patients, 7/8 (87.5%) of which exhibited at least one *de novo* response (Fig 3a). The breadth of responses to neoantigens ranged from 1 to 16, with a median of 2 per patient (Fig 3b). The total magnitude of the neoantigen-specific T cell response, defined as the sum of responses to individual responding neoantigens, was between 17 and 5189 (median = 106) Spots Forming Cell (SFC) per million PBMC (Fig 3c). Of the 5/13 vaccine non-responders 3 had pre-existing neoantigen responses that did not change post vaccination. We were not able to detect an IFN-γ T cell response to any neoantigen in 2/13 patients.

**Figure 3.**
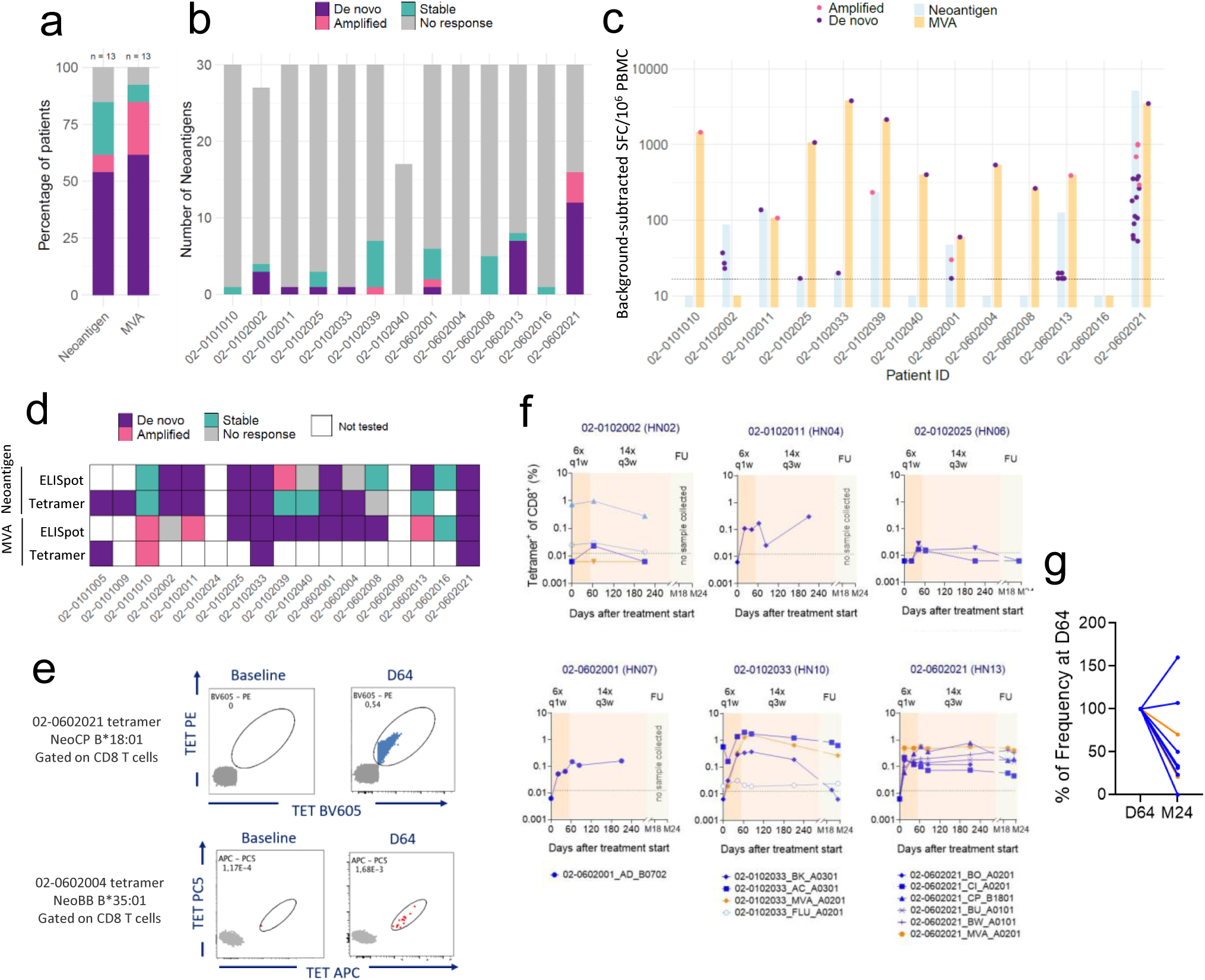
TG4050 induces polyepitopic, long-lasting neoantigen-specific T cell responses. **a, b, c,** T cell responses measured by *ex vivo* IFN-γ ELISpot for 362 individual neoantigens and the MVA peptide pool in 13 Arm A patients at D64 vs baseline. **a**, Frequency of patients with responses to at least one neoantigen and to the MVA peptide pool. **b**, Number of responding neoantigens for each patient within all vaccine neoantigens. **c**, Magnitude of the response to TG4050 treatment (de novo or amplified) for neoantigens (in blue, sum of all responding neoantigens, each represented by one dot) and MVA (in orange). The background-subtracted SFC per million PBMC is reported. **d**, Summary of responses detected by ELISpot and pMHC-I tetramer for neoantigen and MVA at the patient level. **e**, Example of high frequency neoantigen-specific CD8 T cell response detected by both pMHC-I tetramer and ELISpot (patient 02-0602021, upper panel) and of low frequency neoantien-specific CD8+ T cell response detected by pMHC-I tetramer only (lower panel, patient 02-0602004). **f**, Longitudinal pMHC-I tetramer study. Tetramers for MVA and treatment-unrelated viral epitopes (CMV and flu) were used as controls for HLA-A*02 patients. All patients with reactive neoantigen tetramers > 5 / 40,000 CD8+ T cells at at least one time-point are reported. Time-points with frequencies < 5 / 40,000 (0.017% of CD8+ T cells, indicated with a dotted line) are displayed as half this value. **g**, Change in frequency for all reactive tetramers between D64 and M24

*De novo* or amplified T cell responses to the MVA vector backbone were detected in 11/13 patients (84.6%), confirming the ability of the treated patients to mount T cell responses to foreign antigens (Fig 3a). Reflecting the higher number of epitopes included in the peptide pools and/or the xenogeneic nature of the MVA viral sequences, responses to the MVA vector backbone were overall higher in magnitude than those directed to the neoantigens (Fig 3c). The median fold change in SFC per million PBMC between baseline and D64 was 36 (range 3-285) for MVA vs 6 (range 4-33) for neoantigens. Responses to neoantigens were detected in patients with prior exposure to vaccinia, evidenced by the presence of T cell responses to vaccinia antigens at baseline (Supplementary Fig 1). Neither the magnitude of the response to MVA at baseline or D64 nor the TME characteristics (TMB, TME subtype) were associated with the magnitude of the neoantigen-specific response (Supplementary Fig 2).

Neoantigen-specific CD8^+^ T cell responses at baseline and D64 were also analyzed by tetramer in 14 Arm A patients. The number of tetramers tested per patient (range 4 -29) depended on prediction confidence as well as sample and reagent availability. An HLA-A*02:01-restricted tetramer corresponding to a published immunodominant MVA epitope was also included as control for patients carrying this allele. In general, there was good agreement between *ex vivo* IFN-γ ELISpot and tetramer data for both MVA and neoantigen-specific responses (Fig 3d). The tetramer assay detected both high frequency responses to individual neoantigens (example in Fig 3e, upper panel) and neoantigen-specific responses of lower frequency, expected to be below the ELISpot detection level (Fig 3e, lower panel). In line with the ELISpot assay, we observed polyepitopic responses to treatment with a median of 2 (range 1-5) reactive neoepitope-specific tetramers in responding patients. Overall, neoantigen-specific CD8^+^ T cell responses were detected by tetramer in 9/14 (64.3%) patients in Arm A, all of which exhibited at least one *de novo* response to treatment. Taken together, treatment-related T cell responses to vaccine neoantigens were detected by ELISpot and/or tetramer assay in 11/15 (73.3%) Arm A patients with a median of 3 (range 1-16) responding neoantigens per patient (Supplementary Fig 1).

We assessed the dynamics and persistence of the circulating CD8^+^ T cell response in 6 Arm A patients for whom tetramer-reactive cells had been identified and sufficient longitudinal samples were available for analysis (Fig 3f). HLA-A*02:01-restricted tetramers corresponding to MVA, as well as immunodominant CMV and influenza epitopes (representing treatment-unrelated CD8^+^ T cell responses to viral epitopes) were included for patients carrying this allele. Both neoantigen-specific and MVA-specific CD8^+^ T cells were detected as early as two weeks after the first TG4050 dose and increased rapidly during the treatment induction phase. Evaluable neoantigen and MVA specific CD8^+^ T cells with high frequencies at D64 remained detectable after the end of vaccination and up to M24 (one year after the end of TG4050 treatment). The frequencies of control tetramer^+^ cells specific for CMV or influenza epitopes remained unchanged throughout the treatment period. Overall, we observed a gradual but variable decline of tetramer^+^ cells over time and after the end of treatment. The median decrease in neoantigen-specific CD8^+^ T cell frequency over the period of 22 months between D64 and M24 was 3-fold, ranging from a 1.6-fold increase to a 1020-fold decrease (Fig 3g). This decline rate was comparable between MVA- and neoantigen-specific CD8^+^ T cells in the same patient (Fig 3g).

Notably, TG4050 treatment also triggered durable T cell responses when administered at recurrence, as evidenced by the detection of CD8^+^ T cells specific to 2 vaccine neoantigens in one of the 2 Arm B patients treated at relapse (Supplementary Fig 3).

### Characterization of neoantigen-specific T cell responses

Next, we studied the cellular state of the circulating neoantigen-specific CD8^+^ T cells. At D64 (after 7 doses of TG4050), D211 (in the maintenance phase) or M24 (one year after the end of treatment), neoantigen-specific CD8^+^ T cells were mainly effector cells from the effector memory (Tem) or terminally-differentiating effector memory re-expressing CD45RA (Temra) subsets based on the expression of CCR7 and CD45RA on tetramer^+^ cells by flow cytometry (Fig 4a). The distribution between Tem and Temra was dependent on both patient and neoepitope but remained constant for each neoepitope throughout treatment and up to M24, indicating that the effector phenotype was maintained after the end of treatment (Fig 4b and Supplementary Fig 4).

**Figure 4.**
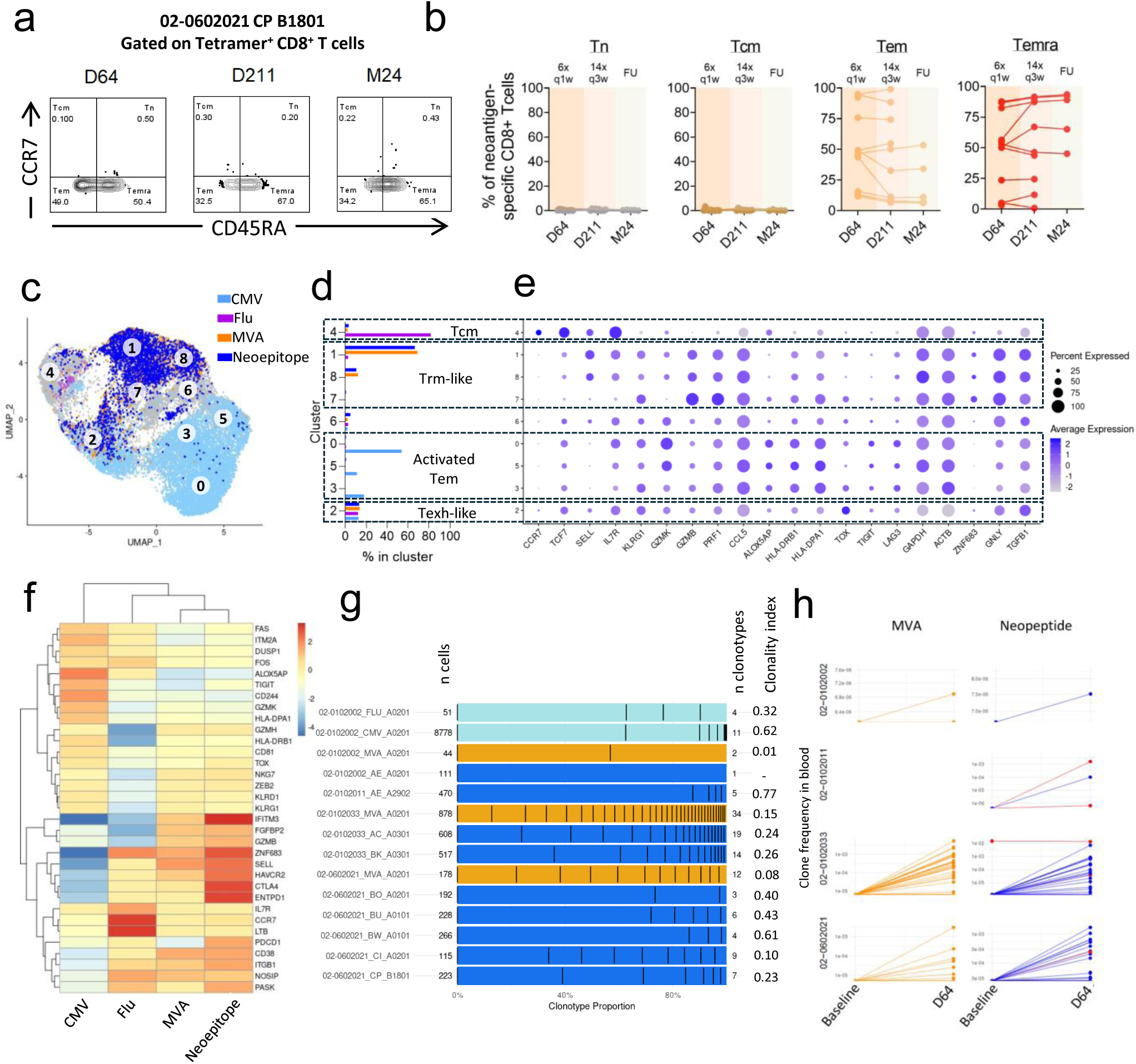
TG4050 induces neoepitope-specific CD8+ T cells with a cytotoxic effector, tissue-resident signature. **a,b.** Phenotyping of neoepitope-specific Tetramer+ CD8+ T cells. **a**, Representative gating of Tn, Tcm, Tem and Temra based on CCR7 and CD45RA expression. **b**, Distribution of phenotype in longitudinal samples for all 12 Tetramer+ populations from 6 patients shown in Figure 3g. One line represents one tetramer. Samples not depicted were not available or below limit of detection. **c to g**, 9 neoepitope-specific Tetramer+ populations from 4 patients as well as 5 virus-specific control Tetramer+ populations (3 MVA, 1 CMV and 1 Flu) were sorted at D64 and analyzed by scRNAseq. **c**, UMAP and cluster analysis with populations overlayed according to antigen specificity. For patient-specific overlay, see Supplementary Figure 5. **d**, Distribution of antigen-specific CD8+ T cells in clusters. **e**, cluster annotation based on expression levels of selected lineage markers. **f**, “Z-score” Gene expression levels between CD8+ T cells specific for CMV, Flu, MVA and neoepitope. A curated gene list based on association with antigen-experienced T cells is shown. **g**, Total number and relative proportion of clonotypes found for each patient and epitope by scRNAseq. **h,** Bulk TCR Vβ DNA sequencing was performed in the blood and in the resected tumor. The evolution of frequency of clones matched to Tetramer+ CD8+ T cells Vβ within all T cell clones in the blood, at baseline and D64, is shown. T cell clones also found in the resected tumor by bulk TCR DNA sequencing are highlighted in red.

We sorted tetramer^+^ cells from 4 Arm A patients at D64 and analyzed their transcriptomic profile and paired T cell receptor (TCR) sequences by single cell RNA sequencing (scRNAseq). Transcriptomic profiles of 24,027 cells were retrieved, and 13,024 T cells were unambiguously assigned to barcodes from individual neoepitope, MVA, CMV or influenza tetramers. Uniform Manifold Approximation and Projection (UMAP) dimensionality reduction and Louvain clustering identified 11 clusters, 9 of which were assigned to CD8^+^ T cell states and selected for further characterization (Fig 4c). The other two clusters showed monocytic and B cell signatures and were considered non-contributive. Consistent with previous reports (Yossef et al, 2023, Schmidt et al, 2023), the majority (82%) of influenza-specific CD8 T cells were found in cluster 4 characterized by a memory quiescent profile with expression of CCR7, TCF7, SELL (CD62L) and ILR7 but relatively low expression of cytotoxic markers such as KLRG1, GZMB, GZMH or PRF1 (Fig 4d, e). Similarly, the majority (83%) of CMV-specific CD8^+^ T cells were found in clusters 0, 5 and 3 with a highly differentiated activated effector signature indicated by expression of cytotoxic markers and activation markers ALOX5AP, HLA-DRB1, HLA-DPA1. Immune checkpoint receptors TIGIT and LAG3 linked to chronic antigen exposure were also enriched in these clusters, in line with repeated antigen stimulation in CMV infection.

Tetramer^+^ cells specific for MVA and neoepitopes showed remarkable transcriptional homogeneity between patients (Supplementary Fig 5). Overall, 81% of MVA-specific and 77% of neoepitope-specific cells were found in clusters 1 and 8, characterized by the expression of the master regulator of tissue-resident memory T cells (Trm) ZNF683 and high expression of cytotoxic markers GNLY, GZMB and PRF1. These cells were enriched in transcripts linked to tissue homing or migratory abilities such as SELL (CD62L), ITGB1 or CX3CR1, suggesting that they have the ability to enter and exit tissues where their target antigens are expressed. Finally, between 12% and 14% of cells for each antigen category were found in cluster 2, mapped to highly differentiated, exhausted (Texh)-like effector cells showing signs of dysfunction through high expression of TOX, a master regulator of exhaustion, and low proliferation through low expression of housekeeping genes GAPDH and ACTB.

Because MVA- and neoepitope-specific CD8^+^ T cells significantly overlapped in their cluster distribution, we investigated whether some transcripts were differentially expressed between the two subsets. We focused on the expression levels of selected transcripts commonly linked to tumor-associated T cells because they mark chronic antigen stimulation. Differential gene expression analysis revealed that compared to MVA-specific CD8^+^ T cells, neoepitope-specific CD8^+^ T cells had higher expression of markers associated with antigen stimulation such as PDCD1 (PD-1, log2FC = 2.8, p = 3E-23), TNFRSF9 (4-1BB, log2FC = 1.5, p = 2E5), CTLA4 (log2 FC = 1.3, p=0.03) or TIGIT (log2FC = 1.3, p = 2E-14) (representative genes shown in Fig 4f). Expression of ENTPD1 (CD39) also tended to be higher in neoepitope-specific CD8 T cells (log2FC = 1.8) but the difference was not statistically significant. GSEA analysis of the differentially expressed genes confirmed that neoepitope-specific CD8^+^ T cells were enriched in pathways linked to chronic antigen stimulation (REACTOME_COSTIMULATION _BY_THE_CD28_FAMILY FDR = 6E-4, REACTOME_PD_1_SIGNALING FDR = 1E-3), whereas MVA-specific CD8^+^ T cells were enriched in pathways linked to translation and metabolism (REACTOME_TRANSLATION FDR = 3E-6).

To analyze neoantigen-specific CD8^+^ T cell clonality, we used the paired TCRA and TCRB sequences obtained by scRNAseq. Between 1 and 19 clonotypes were identified for each neoepitope (median of 6), 2 to 34 for MVA, 4 for influenza and 11 for CMV (Fig 4g). Clonality indexes were calculated to compare the diversity of the response across conditions. As expected from their less differentiated central memory signatures, influenza-specific CD8^+^ T cells had a lower clonality index than CMV-specific cells (0.32 vs 0.62) indicating higher diversity and lower clonal expansion. The clonality index for neoepitopes ranged between 0.10 and 0.77 with a median of 0.33, showing that multiple clones specific for the same neoepitope expand after treatment generally without dominance by a single clone. Together, this demonstrates that the neoantigen-specific response to TG4050 is both polyepitopic and polyclonal within individual epitope reactivities.

We next assessed whether post-treatment CD8^+^ T cell responses originated from the expansion of pre-existing T cell clones infiltrating the tumor before resection. We performed bulk sequencing of the TCR Vβ repertoire from resected tumor samples and blood prior to treatment and blood at D64 after 7 doses of TG4050, identifying clonotypes based on the TCRB CDR3 nucleotide sequence. 43/68 (63%) of the neoepitope-specific clonotypes and 37/48 (77%) of the MVA-specific clonotypes identified by scRNAseq were matched to TCR Vβ sequences found in the blood. Of those, 91% of the neoepitope and 95% of the MVA-specific TCRB expanded between Baseline and D64 (Fig 4h). In 3 of the 4 patients, at least one of the neoepitope-specific clones expanded after treatment was present in the tumor before treatment (Fig 4h, red lines); none of the MVA-specific clones were found in the tumor. In these 4 patients and 3 additional Arm A patients for whom TCR Vβ sequencing could be performed without matching TCR specificity information, we observed that among all T cell clonotypes present in the tumor at the time of surgery, a median of 1.97% (range 0.99 – 2.81%) was expanded in the blood at D64 after TG4050 treatment (Supplementary Figure 6). Together, these findings suggest that TG4050 induces *de novo* CD8^+^ T cell responses against encoded neoantigens as well as expands pre-existing tumor-associated T cell clones.

## Discussion

Here, we show in a randomized Phase I study that an MVA-vectored individualized neoantigen therapeutic vaccine (INTV) may prevent relapse when administered as monotherapy in an adjuvant treatment regimen in high risk resected locally advanced HNSCC patients. Out of 16 patients treated immediately with TG4050, none relapsed after 2 years, whereas 3 of 16 patients randomized to the control arm experienced disease recurrence. Even at this small sample size, these results are encouraging and bring proof-of-principle that TG4050 is immunogenic as monotherapy and may have a role in preventing tumor recurrence.

Our study enrolled patients representative of locally advanced HPV-negative HNSCC patients. The median TMB was 3.02 Mut/Mb, comparable to previous studies in HNSCC (Stransky et al, 2011). The majority of patients harbored immune-deserted tumors with low levels of immune infiltrates. Such tumors are typically associated with lower response rates to immunotherapy due to the combination of medium to low neoantigen abundance and poor immune engagement. Here, we were able to identify candidate immunogenic neoantigens for all patients with sequencing data. Importantly, analysis of the tumor mutanome of the patients included in our study revealed a very low frequency of shared variants. After selection of predicted immunogenic neoantigens, there was no common vaccine target across our study population. Thus, individualized vaccines are a highly relevant addition to the therapeutic options in HNSCC.

TG4050 is expected to work by inducing tumor-specific T cell responses which target and eliminate residual tumor cells after surgery in patients treated immediately after randomization. We find that TG4050 indeed primed and amplified circulating polyepitopic and polyclonal T cell responses to vaccine-encoded neoantigens. T cell responses were detected in the majority (73%) of Arm A patients by *ex vivo* IFN-γ ELISpot and/or tetramer. There were differences between patients in terms of breadth of response, with between 1 and 16 (median = 3) responding neoantigens out of the 30 vaccine-encoded neoantigens, and in terms of magnitude of the response, with between 17 and 5189 (median = 106) SFC per million PBMC. Due to the relatively low sensitivity of *ex vivo* IFN-γ ELISpot (detecting >16 T cells per million PBMC, which can be approximated to >16 T cells per mL of blood) and the technical limitations in screening large numbers of tetramers, it is likely that the number of responding neoantigens is underestimated and that responses at lower frequency, still potentially biologically relevant, remain undetected.

An important observation was that TG4050 expanded both neoantigen-specific CD8^+^ T cell clonotypes that were not detected prior to treatment and clonotypes present in the tumor prior to surgery. The latter suggests that vaccine-encoded neoantigens contain neoepitopes that are both immunogenic and tumor-relevant. At the transcriptomic level, neoepitope-specific CD8^+^ T cells were effector cells closely resembling antiviral CD8^+^ T cells induced against the MVA vector backbone. Compared to circulating influenza-specific and CMV-specific CD8^+^ T cells in the same patients, circulating neoantigen- and MVA-specific CD8^+^ T cells had a higher expression of markers of cytotoxicity and of ZNF683, a crucial regulator of tissue residency. This suggests vaccine-induced neoantigen-specific CD8^+^ T cells have the potential to patrol tissues and kill their target cells. Importantly, T cell responses persisted at least one year after the end of treatment, indicating long-term anti-tumor CD8^+^ T cell responses even after cessation of TG4050 treatment.

Using a viral vector for individualized vaccines has potential drawbacks, such as more complex manufacturing and the pre-existence or induction or anti-vector immunity in the target population. In our study, TG4050 was successfully manufactured for all but 3 eligible patients (92%), supporting the feasibility of the approach. In terms of immunogenicity, differences in analytical methodology make robust comparisons with other platforms difficult. Nonetheless, based on published data we estimate that we observe levels of neoantigen-specific T cell responses with TG4050 monotherapy that are comparable to those published by others with mRNA in combination with ICIs (median of 3 neoantigens per patient for our study vs 2 per patient in Rojas et al, 2023 and Lopez et al, 2025, 4 per patient in Gainor et al, 2024).

We saw no correlation of either breadth of magnitude of neoantigen responses to MVA T cell reactivities, either pre-existing or treatment-induced. While we expect that both T cell and antibody responses to the vector are induced during treatment (based on this study and Di Bisceglie et al. 2014), the fact that we measure polyepitopic and durable neoantigen-specific responses confirms that anti-vector immunity does not overpower the response to TG4050 treatment.

We had anticipated a positive link between anti-vector T cell responses and those against neoantigens. Presentation of neoepitopes in the context of a viral infection provides innate immune stimulation required to shape cytotoxic T cell responses. In addition, the co-expression of strongly immunogenic MVA sequences can provide T-cell help for the response to the MHC class I neoantigens. Even though we did not observe an association between the magnitude of the neoantigen- and MVA-specific responses, the viral vector may still influence the response qualitatively. Immune responses to MVA have been reported to have a strong tissue-resident CD8^+^ T cell component (Rotrosen et al, 2023), a feature critical for protection against the virus through its natural route of transmission but also for anti-tumor immunity. In line with this, our data shows that TG4050-induced neoantigen-specific CD8^+^ T cells express ZNF683, a critical regulator of tissue-residency. Published data demonstrate that MVA induced T cell responses are durable in human beings and that following smallpox vaccination 90% of subjects maintaining substantial cellular immunity 25-75 years after vaccination and T cell responses declining with a half-life of 8-15 years (Hammarlund et al, 2003). Here, we show that TG4050-induced CD8^+^ T cell responses can persist at high levels up to one year after the end of treatment, with similar kinetics for CD8^+^ T cells reactive with MVA peptides and neoantigens, thus suggesting that neoantigen responses may be equally durable to those induced by the vector backbone. This would have important implications for long-term protection against tumor recurrence from TG4050.

Two patients from Arm B were treated with TG4050 at recurrence. In one patient, a small number of neoantigen tetramers was tested and no response was found. In the other patient, tetramer^+^ cells were detected for 2 neoantigens and persisted up to the last time-point tested (D158). In spite of this documented immune response the patient did not achieve a measurable clinical response, suggesting that for clinical success in advanced disease additional strategies are needed. We consider that adjuvant use is the most promising window of opportunity for cancer vaccines, when the neoantigen-specific T cells induced can target micrometastatic disease in tissues before the development of an immunosuppressive environment. Our data suggest that in this setting, monotherapy may be sufficient. On the other end, in settings where the tumor has already developed mechanisms to evade T cell-mediated killing, combinations may be required to improve patients outcome.

Two randomized phase 3 trials in locally advanced HNSCC now support the use of anti-PD1 antibodies in the peri-operative and adjuvant settings, respectively. There is a strong clinical and scientific rationale for a synergy between a neoantigen targeting vaccine and ICIs, including the expression of PDCD1 in the neoantigen-specific CD8^+^ T cells observed in our scRNAseq data. As ICI efficacy is mediated by the re-invigoration of naturally occurring T cells in the cancer tissue, benefit from ICI will be limited if tumors have low numbers of naturally existing tumor-reactive T cells. Combination with TG4050 could prime neoantigen-specific *de novo* responses and amplify pre-existing ones and thus be an ideal backbone going forward in advanced disease.

## Methods

### Study design

TG4050.02 is a phase 1/2 multicenter, open-label, randomized trial of adjuvant personalized cancer vaccine TG4050 in HNSCC (NCT04183166).

The trial was conducted in accordance with the Declaration of Helsinki, International Conference on Harmonization E6 guidelines and Good Clinical Practice guidelines. The protocol and all amendments were approved by the appropriate ethics body at each participating site. All the participants provided written informed consent.

The primary objective of the phase 1 was to evaluate the safety and tolerability of multiple subcutaneous (SC) injections of a mutanome-directed active immunotherapy (TG4050) in participants with resectable, locoregionally advanced HNSCC initiated at completion of primary treatment or at the time of recurrence. Secondary objectives included the evaluation of efficacy in terms of Disease-Free-Survival (DFS) after completion of primary treatment and tumor response and progression free survival according to RECIST 1.1 in relapsing participants, as well as the rate of failure to provide TG4050. Exploratory objectives included the assessment of tumor mutational burden (TMB) and neoantigen frequency at Baseline and at recurrence, changes in circulating immune cells, induction of tumor specific T cells, T cell changes in tumor immune infiltrate and environment at baseline and at recurrence and quantification of tumor and immune-related biomarkers. Eligible participants were 18 years of age or older and had newly diagnosed, locally advanced, HPV negative, resectable stage III or IV oropharyngeal, laryngeal, hypopharyngeal, or oral cavity HNSCC. Participants had to have an Eastern Cooperative Oncology Group performance-status score of 0 or 1 and had to be eligible for primary surgery and adjuvant radiotherapy (RT) combined or not with cisplatin.

Upon confirmation of complete response by imaging 3 months after completion of primary treatment, participants were randomized to receive immediate vaccination with weekly subcutaneous doses of TG4050 for 6 weeks followed by a maintenance period of one dose every 3 weeks for up to 20 doses (Arm A) or to a delayed vaccination arm with the same vaccination regimen initiated at relapse in combination with SOC (Arm B). Participants in Arm A could be given a second series of TG4050 in combination with SOC at relapse. Randomization was performed based on a dynamic method (minimization) according to pathological tumor stage (pIII vs. pIV), cisplatin use, and center.

### Safety

Safety analyses were performed in the safety population defined as all randomized patients who received at least one injection of TG4050. AEs were reported per National Cancer Institute Common Terminology Criteria for Adverse Events (CTCAE), v.5.0 for 30 days after the last dose of TG4050 or standard of care. Safety was assessed through summaries of DLTs, AEs, changes in laboratory test results, and changes in other safety assessments as physical examinations including ECOG, vital signs and ECGs. A safety run-in phase was conducted in the first 3 participants in each arm treated for 6 weeks. During this period, DLT was defined as any death or any treatment-emergent AE grade ≥3 toxicity at least possibly related to TG4050, with the exception of pre-existing grade 2 fatigue at baseline that would worsen up to grade 3. Beyond the safety run-in phase, DLT was defined as any treatment-emergent AE grade ≥3 toxicity at least possibly related to TG4050, with the exception of pre-existing grade 2 fatigue at baseline that would worsen up to grade 3. Regular safety assessments as prespecified in the clinical trial protocol were performed by an external, independent data safety monitoring committee.

### Efficacy

Analysis for efficacy in terms of DFS was performed in the per protocol population defined for Arm A as a minimum exposure of 6 doses of TG4050 or for Arm B a 6-week medical follow-up without relapse. Efficacy was assessed as per standard medical practice which included physical examination of the head and neck including panendoscopy if appropriate, MRI and/or CT of the head and neck and/or PET CT in the event of clinical signs or symptoms of recurrence every 12 weeks during the first 24 months after randomization and every 24 weeks thereafter. Tumor response and progression free survival in relapsing participants was evaluated by CT or MRI of the head and neck, and CT of the chest, abdomen and pelvis every 6 weeks until documented progression (not presented due to the low number of evaluable patients at the cut-off date).

### Patient tumor and tissue sequencing

Patient tumor samples were obtained from resection material and immediately frozen or formalin-fixed and paraffin embedded (FFPE). Blood specimens were collected in PAXgene tubes. Whole exome sequencing (WES) was performed on blood and tumor samples (preferably frozen) using the Illumina Nextera Rapid Capture Exome v1.2. RNA sequencing was performed on tumor samples only. Libraries from total RNA with low degradation profiles (RIN>6), were prepared using Illumina’s TruSeq RNA Access Library Prep kit. Exome capture was performed for RNA material with more degradation (RIN>1 & DV200>60%). Highly degraded RNA samples were replaced by backup samples when available. All sequencing was performed by Fulgent Genetics (Temple City, CA, USA) on Illumina HiSeq in 2x150bp paired mode.

Tumor profiling was performed using BostonGene Tumor Portrait™. The TME subtype was determined according to the categorization strategy described in Bagaev et al, 2021.

### Mutation identification and neoantigen selection

Single nucleotide variants (SNVs) and small insertions/deletions (indels) in the tumor genome were identified and characterized using a consensus-based strategy that integrates different variant callers on tumor – normal paired WES data. Expression of the identified somatic variants was assessed using RNA-seq data, and only expressed variants were selected for subsequent neoantigen prediction.

The immunogenic potential of mutated peptides derived from tumor-specific alterations was evaluated using a modular neoantigen prediction pipeline developed by NEC Bio, which integrates proprietary machine learning (ML) algorithms for T cell epitope prediction. For class I, neoantigen predictions were performed for HLA-A and HLA-B alleles in each patient, considering peptides of 9 and 10 amino acids in length. For class II, predictions were conducted for HLA-DR alleles, considering 15-mers. The pipeline integrates multiple determinants of epitope immunogenicity, including: (i) expression level of the mutated peptide; (ii) probability of antigen processing and presentation, the pipeline models the full antigen-processing cascade (including proteasomal cleavage and TAP transport for class I, and endosomal proteolysis with HLA-DM – mediated peptide exchange for class II) to estimate the likelihood that each mutated peptide is generated, bound to its cognate MHC molecule, and stably presented on the tumor cell surface (iii) binding affinity to the patient-specific HLA alleles predicted using binding assays; and (iv) likelihood of T cell receptor recognition predicted based on an immunogenicity model computing a sequence similarity – based distance metric quantifying each peptide’s distinctiveness from self-proteins.

### Design of vaccine cassettes

For each neopeptide, a ∼29 amino acid window with the mutation in central position was selected. Candidate neopeptides were ranked based on the highest composite immunogenicity scores and used for the design of the personalized vaccine cassettes by VacDesignR®, a proprietary vaccine design software developed by Transgene SA. Up to 30 neopeptides with no predicted transmembrane domain, low hydrophobicity and no homologous domains between peptides were selected and optimally assembled in up to three fusion proteins of up to 10 neopeptides separated by GSG, GAS or GTS linkers. Each fusion protein was preceded by a signal peptide derived either from the rabies glycoprotein or the measles F glycoprotein. Expression of the three fusion constructs is driven by distinct vaccinia virus promoters: pH5R, pC11R, and p7.5K.

### TG4050 manufacturing

Synthetic genes encoding the neoepitope polypeptides and their respective promoters were synthesized by ThermoFischer Scientific (Regensburg, Germany), with codon optimization for human expression. Specific sequence motifs, such as 5TNT and homopolymer stretches longer than 5 base pairs, were excluded to improve mRNA stability and expression.

The DNA fragments corresponding to the three expression cassettes were cloned into a vaccinia shuttle plasmid between 500 bp regions homologous to the MVA 164R and 165R genes, enabling targeted insertion into deletion III of the MVA genome via homologous recombination (Supplementary Figure 7). Recombinant MVAs were generated in primary chicken embryo fibroblasts (CEFs). Cells were infected with a parental MVA vector containing an mCherry reporter gene in deletion III and subsequently transfected with the recombinant shuttle plasmid. Recombinant clones lacking fluorescence were selected by plaque purification. The integrity and identity of the expression cassettes were confirmed by sequencing PCR amplicons. The MVA strain originates from Dr. Mayr and is derived from MVA-575 (Mayr et al, 1975).

Clinical-grade vaccine batches were produced aseptically under good manufacturing practices conditions using CEF as a cell substrate. Following patient specific recombinant MVA amplification on host cells, the viral particles are extracted mechanically, purified and formulated using tangential flow filtration. The identity of the neopepitope expression cassettes were confirmed before shipment for patient administration. Vaccine batches were stored at -20 °C.

### Ex vivo IFN-γ ELISpot

For each neoantigen 300,000 cryopreserved PBMCs were seeded in triplicate wells in TexMACS medium (Miltenyi) in Fluorospot plates (Mabtech FSP-01A-10) and stimulated for 16-24h with 10 μg/ml of a custom-made pool of 3 15-mers overlapping by 11aa covering the neoantigen sequence (Proteogenix). A concentration of DMSO equivalent to that in the peptide-stimulated conditions was used as a negative control and anti-CD3 as a positive control. A mix of Vaccinia CD8 and CD4 immunodominant epitopes (Mabtech 3670-1 and 3635-1) was used to assess anti-MVA responses. The CEFX Ultra SuperStim Pool (JPT PM-CEFX-1) was used as a treatment-unrelated antigen control. Anti-CD3 was used as a sample quality control. Plates were revealed according to the manufacturer’s protocol and spots per well (SFC) counted using a MultiSpot Spectrum MSR08 with fluorescent lamp iSpot (AID).

T cell responses were analyzed by distribution-free resampling (DFReq) as described in (Moodie et al, 2010). For each neoantigen, a response was reported if the DFReq test between peptide-stimulated and matching unstimulated control as well as the DFReq test between peptide-stimulated at baseline and D64 were positive, and if at least 5 SFC per well were counted at D64. T cell responses were defined as de novo if not detected at baseline but detected at D64, amplified if detected at baseline but increased more than 2-fold at D64, and stable if detected at baseline and D64 at comparable levels (less than 2-fold increase). Only de novo and amplified responses were considered as responses to treatment.

### Peptide-MHC class I tetramers

Biotinylated HLA-Class I monomers (easYmer) were combined with each 9-mer peptide and labeled with 2 different fluorochromes conjugated streptavidins. Immunodominant viral peptides restricted to HLA-A*02:01 were used as control: FLU MP 58-66 (GILGFVFTL), CMV pp65 495-503 (NLVPMVATV) and MVA_KV-9 (KVDDTFYYV).

CD8+ T cell enrichment was performed from cryopreserved PBMCs using the EasySep Negative Human CD8^+^ Enrichment Kit (StemCell). Enriched CD8^+^ T cells were stained with a viability stain (SYTOX Blue, Invitrogen S34857), the respective tetramer mix and T cell phenotypic markers CD3 BUV737 (BD 12751), CD8 BUV396 (BD 563795), CD45RA FITC (BD 555488), CCR7 BV421 (Biolegend 353208), CD27 PC7 (BD 560609), PD-1 APC-CY7 (Biolegend 329922) and CD39 Percp-ef710 (eBioscience 46-0399-42). Samples were acquired on a BD Fortessa cytometer and analyzed using FlowJo v10.10.0.

Tetramer-positive populations were analyzed by combinatorial gating within live CD8^+^ T cells. For screening of T cell responses at baseline vs D64, where at least 200,000 CD8^+^ T cells were acquired for each sample, a tetramer response was reported if a distinct population of at least 10 cells positive for both tetramer fluorophores was present. Tetramer responses were defined as de novo if not detected at baseline but detected at D64, amplified if detected at baseline but increased more than four-fold at D64, and stable if detected at baseline and D64 at comparable levels (less than four-fold increase). Only *de novo* and amplified responses were considered as responses to treatment.

For longitudinal studies, where a lower number of CD8^+^ T cells were available for analysis, a threshold was applied to enable inter-sample comparisons. Samples with > 40,000 CD8^+^ T cells were included in the analysis and a minimum of 5 cells positive for both tetramer fluorophores were required to report a response. Samples with frequencies < 5/40,000 (0.017% of CD8^+^ T cells) were considered below detection limit and imputed as half this value.

### scRNAseq

Barcoded tetramers were prepared following the same method described above with the exception that streptavidin was conjugated to a TotalSeq-C DNA barcode (Biolegend) and PE. A second non-barcoded tetramer was prepared with a different fluorochrome and staining of enriched CD8 T cells was performed as described. Tetramer positive cells at D64 were sorted using a BD ARIA Fusion FACS and mixed with tetramer negative CD8^+^ and CD4^+^ T cells in the same proportion. The captured cells were subjected to scRNAseq with paired sequencing of the TCRA and TCRBCDR3 regions. Libraries were generated with the 10X Chromium 5’ GE kit. Library for gene expression was sequenced by NovaSeq (Illumina) aligned to GRCh38-2020-A genome. Library for VDJ TCR was sequenced by NextSeq2000 (Illumina) and aligned to vdj_GRCh38_alts_ensembl-3.1.0-3.1.0. Cellranger-8.0.1 was used to process raw reads and downstream analysis was performed using the Seurat R package (Butler et al, 2018). The following filters were used to discard doublets, debris and dying cells: 1000 < nFeature_RNA < 35000, percent of mitochondrial genes < 10 and 1600 < nCount_RNA < 35000. Only cells with complete TCR information (exactly 1 TRA and 1 TRB sequence) were kept. Cells were demultiplexed based on tetramer barcodes with the MULTIseqDemux function (quantile 0.7). Negative and doublet cells were qualified as not assigned. The clustering was performed using the Louvain algorithm (resolution 0.3), excluding TCR genes to not drive the clustering. Clusters were annotated manually based on lists of published circulating T cell subset markers (Yossef et al, 2023, Schmidt et al, 2023, Rojas et al, 2023). Differential gene expression analysis was performed using the FindMarkers function on single cell data. Clonality of barcoded cells was determined based on the unique paired TCRA and TCRB CDR3 nucleotide sequence and their v-usage. TCRA and TCRB pairs with more than 5 cells were considered one clonotype.

### Bulk TCR Vβ sequencing

Bulk TCR sequencing was performed on DNA and RNA extracted from tumor tissue at surgery and PBMCs collected at Baseline and D64. DNA libraries (2 x100bp) were prepared with AmpliSeq^TM^ TCR beta-SR Panel (Illumina). Demultiplexing was performed with Illumina bcl2fastq (version2.20), adapters were trimmed with Skewer (version 0.2.2) and overlapping paired reads were merged into single reads before reconstruction of the CDR3β regions using NGmerge. Reconstruction of T cell receptor sequences was performed using RTCR (Gerritsen et al. 2016). Clone counts were determined for each unique CDR3 nucleotide (CDR3.nt) sequence at each time point and sample, respectively. Clone proportions for given T cell clones were calculated relative to the total number of detected clones per sample. Tumor-infiltrating lymphocytes (TILs) were defined as clones detected in the tumor sample (in either DNA- or RNA sequencing). Clones not specific to the respective patient, i.e. shared by multiple patients, were removed before downstream analyses.

To identify significantly expanded or contracted TIL clones, Fisher’s exact test was applied to compare clone counts between baseline and D64 samples for each patient. Clones exhibiting an absolute fold-change ≥ 3 and a corresponding *p*-value ≤ 0.001 were considered statistically significant.

To validate TCR clonotypes identified in the bulk sequencing dataset, amino acid sequences of TRB CDR3 regions were compared between the scRNA-seq and bulk data. Two sequences from bulk and scRNA-seq were considered matching if either sequence was contained within the other or identical. Those sequences were classified as validated by scRNA-seq.

### Statistical analyses

Descriptive statistics were used to summarize patient demographics and clinical characteristics and safety data. Disease-free survival (DFS) was defined as the time from randomization to the first occurrence of locoregional recurrence, distant metastasis, second primary squamous cell carcinoma of the head and neck (SCCHN), or death from any cause, whichever occurred first. DFS was estimated using Kaplan-Meier method, with comparisons between the 2 arms performed using a one-sided log-rank test. A p-value of 0.05 was considered statistically significant. No formal sample size calculation was performed for this phase I First-in-human study.

Statistical analyses were conducted using SAS software version 9.4.

## Data Availability

All data produced in the present study are available upon reasonable request to the authors

## Acknowledgements

We wish to thank patients and their families for participating in the study. We are grateful to medical and administrative staff at the study sites, Transgene and NEC Bio for their support. Collaborators at Fulgent Genetics (El Monte, CA), Quality Assistance (Thuin, Belgium) and Cegat (Tübingen, Germany) provided technical and analytical support. Funding was provided by bpifrance, Transgene and NEC Bio.

## Authors contributions

C. Ottensmeier, K. Bendjama, M. Brandely, E. Quemeneur, C. Le Tourneau, A. Tavernaro, developed the concept and designed the clinical trial. E. Piaggio, K. Bendjama secured funding. A. Tavernaro, G. Lacoste, E. Dochy, M. Brandely oversaw the clinical trial as sponsor representatives. J-P. Delord, C. Ottensmeier and C. Le Tourneau recruited patients. T. Jones, A. Schache participated in patient recruitment and performed surgeries. V. Schoettel was the pharmacovigilance physician. B. Bastien performed statistical analyses. N. Silvestre, J.B Marchand, S. Robin designed the MVA construct and production process. B. Grellier, P. Brattas, K. Onoguchi, Y. Yamashita, H. Fontenelle designed neoantigen vaccines. O. Lantz, A. Lalanne, K. Bendjama designed the translational strategy. K. Bidet Huang, C. Spring-Guisti designed and coordinated ELISpot analyses. A. Lalanne, C. Jamet performed tetramer and scRNAseq experiments. A-L. Le Gac, J. Deforges performed bioinformatics analyses for scRNAseq. M. Eggert Martinez, O. Baker performed bulk TCR analysis. A. Lalanne, C. Jamet, K. Bendjama, O. Lantz, K. Bidet Huang analyzed and interpreted translational data. K. Bidet Huang, E. Dochy wrote the manuscript with critical discussion and review from all authors. M. Ceppi, A. Riva oversaw departments involved at Transgene

## References

1. Bray F., Laversanne M., Sung H., Ferlay J., Siegel RL, Soerjomataram I., Jema A., Global cancer statistics 2022: GLOBOCAN estimates of incidence and mortality worldwide for 36 cancers in 185 countries; CA Cancer J Clin 2024;74:229–263; doi: 10.3322/caac.21834

2. Psyrri A, Rampias T, Vermorken JB: The current and future impact of human papillomavirus on treatment of squamous cell carcinoma of the head and neck. Ann Oncol 25:2101–2115, 2014

3. Machiels JP, René Leemans C, Golusinski W, Grau C, Licitra L, Gregoire V. Squamous cell carcinoma of the oral cavity, larynx, oropharynx and hypopharynx: EHNS-ESMO-ESTRO Clinical Practice Guidelines for diagnosis, treatment and follow-up. Ann Oncol 2020; 31:1462–75.

4. National Comprehensive Cancer Network. NCCN Clinical Practice Guidelines in Oncology (NCCN Guidelines): Head and Neck Cancers, version 2.2023. 2023. https://www.nccn.org/professionals/physician_gls/pdf/head-and-neck.pdf (accessed Aug 1, 2023).

5. Haque S, Karivedu V, Riaz MK, Choi D, Roof L, Hassan SZ, Zhu Z, Jandarov R, Takiar V, Tang A, Wise-Draper T. High-risk pathological features at the time of salvage surgery predict poor survival after definitive therapy in patients with head and neck squamous cell carcinoma. Oral Oncol. 2019 Jan;88:9–15. doi: 10.1016/j.oraloncology.2018.11.010. Epub 2018 Nov 16. PMID: 30616803; PMCID: PMC6327963.

6. Basyuni S, Nugent G, Ferro A, Barker E, Reddin I, Jones O, Lechner M, O’Leary B, Jones T, Masterson L, Fenton T, Schache A. Value of p53 sequencing in the prognostication of head and neck cancer: a systematic review and meta-analysis. Sci Rep. 2022 Dec 1;12(1):20776. doi: 10.1038/s41598-022-25291-2. PMID: 36456616; PMCID: PMC9715723.

7. Nair S, Bonner JA, Bredel M. *EGFR* Mutations in Head and Neck Squamous Cell Carcinoma. Int J Mol Sci. 2022 Mar 30;23(7):3818. doi: 10.3390/ijms23073818. PMID: 35409179; PMCID: PMC8999014.

8. Ferris RL, Blumenschein G Jr, Fayette J, Guigay J, Colevas AD, Licitra L, Harrington K, Kasper S, Vokes EE, Even C, Worden F, Saba NF, Iglesias Docampo LC, Haddad R, Rordorf T, Kiyota N, Tahara M, Monga M, Lynch M, Geese WJ, Kopit J, Shaw JW, Gillison ML. Nivolumab for Recurrent Squamous-Cell Carcinoma of the Head and Neck. N Engl J Med. 2016 Nov 10;375(19):1856–1867. doi: 10.1056/NEJMoa1602252. Epub 2016 Oct 8. PMID: 27718784; PMCID: PMC5564292.

9. Burtness B, Harrington KJ, Greil R, Soulières D, Tahara M et al, Pembrolizumab alone or with chemotherapy versus cetuximab with chemotherapy for recurrent or metastatic squamous cell carcinoma of the head and neck (KEYNOTE-048): a randomised, open-label, phase 3 study, Lancet, 2019; 394:1915–1928. doi: 10.1016/S0140-6736(19)32591-7

10. Uppaluri R, Haddad RI, Tao Y, Le Tourneau C, Lee NY, Westra W, Chernock R, Tahara M, Harrington KJ, Klochikhin AL, Braña I, Vasconcelos Alves G, Hughes BGM, Oliva M, Pinto Figueiredo Lima I, Ueda T, Rutkowski T, Schroeder U, Mauz PS, Fuereder T, Laban S, Oridate N, Popovtzer A, Mach N, Korobko Y, Costa DA, Hooda-Nehra A, Rodriguez CP, Bell RB, Manschot C, Benjamin K, Gumuscu B, Adkins D; KEYNOTE-689 Investigators. Neoadjuvant and Adjuvant Pembrolizumab in Locally Advanced Head and Neck Cancer. N Engl J Med. 2025 Jul 3;393(1):37–50. doi: 10.1056/NEJMoa2415434. Epub 2025 Jun 18. PMID: 40532178.

11. Bourhis J, Auperin A, Borel C, et al: NIVOPOSTOP (GORTEC 2018-01): A phase III randomized trial of adjuvant nivolumab added to radiochemotherapy in patients with resected head and neck squamous cell carcinoma at high risk of relapse. 2025 ASCO Annual Meeting. Abstract LBA2. Presented June 1, 2025

12. Johnson DE, Burtness B, Leemans CR, Lui VWY, Bauman JE, Grandis JR. Head and neck squamous cell carcinoma. Nat Rev Dis Primers. 2020 Nov 26;6(1):92. doi: 10.1038/s41572-020-00224-3. Erratum in: Nat Rev Dis Primers. 2023 Jan 19;9(1):4. doi: 10.1038/s41572-023-00418-5. PMID: 33243986; PMCID: PMC7944998.

13. Mito I, Takahashi H, Kawabata-Iwakawa R, Ida S, Tada H, Chikamatsu K. Comprehensive analysis of immune cell enrichment in the tumor microenvironment of head and neck squamous cell carcinoma. Sci Rep. 2021 Aug 9;11(1):16134. doi: 10.1038/s41598-021-95718-9. PMID: 34373557; PMCID: PMC8352955.

14. Torri M, Sandell A, Al-Samadi A. The prognostic value of tumor-infiltrating lymphocytes in head and neck squamous cell carcinoma: A systematic review and meta-analysis. Biomed Pharmacother. 2024 Nov;180:117544. doi: 10.1016/j.biopha.2024.117544. Epub 2024 Oct 17. PMID: 39418961.

15. Paschold L, Schultheiss C, Schmidt-Barbo P, Klinghammer K, Hahn D, Tometten M, Schafhausen P, Blaurock M, Brandt A, Westgaard I, Kowoll S, Stein A, Hinke A, Binder M. Inflammation and limited adaptive immunity predict worse outcomes on immunotherapy in head and neck cancer. NPJ Precis Oncol. 2025 Aug 5;9(1):272. doi: 10.1038/s41698-025-01020-6. PMID: 40764401; PMCID: PMC12325939.

16. Wang X, Li T, Slebos RJC, Chaudhary R, Guevara-Patino JA, Bonomi M, Saba NF, Chung CH. Clinical significance of peripheral T-cell repertoire in head and neck squamous cell carcinoma treated with cetuximab and nivolumab. Cancer Immunol Immunother. 2025 Mar 8;74(4):142. doi: 10.1007/s00262-025-03993-6. PMID: 40056190; PMCID: PMC11890688.

17. Lai J, Huang R, Huang J. Predicting head and neck cancer response to radiotherapy with a chemokine-based model. Sci Rep. 2025 Aug 4;15(1):28450. doi: 10.1038/s41598-025-13346-z. PMID: 40760079; PMCID: PMC12322276.

18. Zhao M, Schoenfeld JD, Egloff AM, Hanna GJ, Haddad RI, Adkins DR, Uppaluri R. T cell dynamics with neoadjuvant immunotherapy in head and neck cancer. Nat Rev Clin Oncol. 2025 Feb;22(2):83–94. doi: 10.1038/s41571-024-00969-w. Epub 2024 Dec 10. PMID: 39658611.

19. Xie N, Shen G, Gao W, Huang Z, Huang C, Fu L. Neoantigens: promising targets for cancer therapy. Signal Transduct Target Ther. 2023 Jan 6;8(1):9. doi: 10.1038/s41392-022-01270-x. PMID: 36604431; PMCID: PMC9816309.

20. Bendjama K, Quemeneur E. Modified Vaccinia virus Ankara-based vaccines in the era of personalized immunotherapy of cancer. Hum Vaccin Immunother. 2017 Sep 2;13(9):1997–2003. doi: 10.1080/21645515.2017.1334746. Epub 2017 Aug 28. PMID: 28846477; PMCID: PMC5612284.

21. Bagaev A, Kotlov N, Nomie K, Svekolkin V, Gafurov A, Isaeva O, Osokin N, Kozlov I, Frenkel F, Gancharova O, Almog N, Tsiper M, Ataullakhanov R, Fowler N. Conserved pan-cancer microenvironment subtypes predict response to immunotherapy. Cancer Cell. 2021 Jun 14;39(6):845–865.e7. doi: 10.1016/j.ccell.2021.04.014. Epub 2021 May 20. PMID: 34019806.

22. Anzar I, Malone B, Samarakoon P, Vardaxis I, Simovski B, Fontenelle H, Meza-Zepeda LA, Stratford R, Keung EZ, Burgess M, Tawbi HA, Myklebost O, Clancy T. The interplay between neoantigens and immune cells in sarcomas treated with checkpoint inhibition. Front Immunol. 2023 Sep 20;14:1226445. doi: 10.3389/fimmu.2023.1226445. PMID: 37799721; PMCID: PMC10548483.

23. Grellier et al, "VacDesignR®: a computational tool to optimize viral-based individualized neoantigen therapeutic vaccine production", ESMO AI & Digital Oncology 2025

24. Yossef R, Krishna S, Sindiri S, Lowery FJ, Copeland AR, Gartner JJ, Parkhurst MR, Parikh NB, Hitscherich KJ, Levi ST, Chatani PD, Zacharakis N, Levin N, Vale NR, Nah SK, Dinerman A, Hill VK, Ray S, Bera A, Levy L, Jia L, Kelly MC, Goff SL, Robbins PF, Rosenberg SA. Phenotypic signatures of circulating neoantigen-reactive CD8^+^ T cells in patients with metastatic cancers. Cancer Cell. 2023 Dec 11;41(12):2154–2165.e5. doi: 10.1016/j.ccell.2023.11.005. Epub 2023 Nov 30. PMID: 38039963; PMCID: PMC10843665.

25. Schmidt F, Fields HF, Purwanti Y, Milojkovic A, Salim S, Wu KX, Simoni Y, Vitiello A, MacLeod DT, Nardin A, Newell EW, Fink K, Wilm A, Fehlings M. In-depth analysis of human virus-specific CD8^+^ T cells delineates unique phenotypic signatures for T cell specificity prediction. Cell Rep. 2023 Oct 31;42(10):113250. doi: 10.1016/j.celrep.2023.113250. Epub 2023 Oct 12. PMID: 37837618.

26. Stransky N, Egloff AM, Tward AD, Kostic AD, Cibulskis K, Sivachenko A, Kryukov GV, Lawrence MS, Sougnez C, McKenna A, Shefler E, Ramos AH, Stojanov P, Carter SL, Voet D, Cortés ML, Auclair D, Berger MF, Saksena G, Guiducci C, Onofrio RC, Parkin M, Romkes M, Weissfeld JL, Seethala RR, Wang L, Rangel-Escareño C, Fernandez-Lopez JC, Hidalgo-Miranda A, Melendez-Zajgla J, Winckler W, Ardlie K, Gabriel SB, Meyerson M, Lander ES, Getz G, Golub TR, Garraway LA, Grandis JR. The mutational landscape of head and neck squamous cell carcinoma. Science. 2011 Aug 26;333(6046):1157–60. doi: 10.1126/science.1208130. Epub 2011 Jul 28. PMID: 21798893; PMCID: PMC3415217.

27. Rojas LA, Sethna Z, Soares KC, Olcese C, Pang N, Patterson E, Lihm J, Ceglia N, Guasp P, Chu A, Yu R, Chandra AK, Waters T, Ruan J, Amisaki M, Zebboudj A, Odgerel Z, Payne G, Derhovanessian E, Müller F, Rhee I, Yadav M, Dobrin A, Sadelain M, Łuksza M, Cohen N, Tang L, Basturk O, Gönen M, Katz S, Do RK, Epstein AS, Momtaz P, Park W, Sugarman R, Varghese AM, Won E, Desai A, Wei AC, D’Angelica MI, Kingham TP, Mellman I, Merghoub T, Wolchok JD, Sahin U, Türeci Ö, Greenbaum BD, Jarnagin WR, Drebin J, O’Reilly EM, Balachandran VP. Personalized RNA neoantigen vaccines stimulate T cells in pancreatic cancer. Nature. 2023 Jun;618(7963):144–150. doi: 10.1038/s41586-023-06063-y. Epub 2023 May 10. PMID: 37165196; PMCID: PMC10171177.

28. Gainor JF, Patel MR, Weber JS, Gutierrez M, Bauman JE, Clarke JM, Julian R, Scott AJ, Geiger JL, Kirtane K, Robert-Tissot C, Coder B, Tasneem M, Sun J, Zheng W, Gerbereux L, Laino A, Porichis F, Pollard JR, Hou P, Sehgal V, Chen X, Morrissey M, Daghestani HN, Feldman I, Srinivasan L, Frederick JP, Brown M, Aanur P, Meehan R, Burris HA 3rd. T-cell Responses to Individualized Neoantigen Therapy mRNA-4157 (V940) Alone or in Combination with Pembrolizumab in the Phase 1 KEYNOTE-603 Study. Cancer Discov. 2024 Nov 1;14(11):2209–2223. doi: 10.1158/2159-8290.CD-24-0158. PMID: 39115419.

29. Lopez J, Powles T, Braiteh F, Siu LL, LoRusso P, Friedman CF, Balmanoukian AS, Gordon M, Yachnin J, Rottey S, Karydis I, Fisher GA, Schmidt M, Schuler M, Sullivan RJ, Burris HA, Galvao V, Henick BS, Dirix L, Jaeger D, Ott PA, Wong KM, Jerusalem G, Schiza A, Fong L, Steeghs N, Leidner RS, Rittmeyer A, Laurie SA, Gort E, Aljumaily R, Melero I, Sabado RL, Rhee I, Mancuso MR, Muller L, Fine GD, Yadav M, Kim L, Leveque VJP, Robert A, Darwish M, Qi T, Zhu J, Zhang J, Twomey P, Rao GK, Low DW, Petry C, Lo AA, Schartner JM, Delamarre L, Mellman I, Löwer M, Müller F, Derhovanessian E, Cortini A, Manning L, Maurus D, Brachtendorf S, Lörks V, Omokoko T, Godehardt E, Becker D, Hawner C, Wallrapp C, Albrecht C, Kröner C, Tadmor AD, Diekmann J, Vormehr M, Jork A, Paruzynski A, Lang M, Blake J, Hennig O, Kuhn AN, Sahin U, Türeci Ö, Camidge DR. Autogene cevumeran with or without atezolizumab in advanced solid tumors: a phase 1 trial. Nat Med. 2025 Jan;31(1):152–164. doi: 10.1038/s41591-024-03334-7. Epub 2025 Jan 6. PMID: 39762422; PMCID: PMC11750724.

30. Di Bisceglie AM, Janczweska-Kazek E, Habersetzer F, Mazur W, Stanciu C, Carreno V, Tanasescu C, Flisiak R, Romero-Gomez M, Fich A, Bataille V, Toh ML, Hennequi M, Zerr P, Honnet G, Inchauspé G, Agathon D, Limacher JM, Wedemeyer H. Efficacy of immunotherapy with TG4040, peg-interferon, and ribavirin in a Phase 2 study of patients with chronic HCV infection. Gastroenterology. 2014 Jul;147(1):119–131.e3. doi: 10.1053/j.gastro.2014.03.007. Epub 2014 Mar 18. PMID: 24657484.

31. Rotrosen E, Kupper TS. Assessing the generation of tissue resident memory T cells by vaccines. Nat Rev Immunol. 2023 Oct;23(10):655–665. doi: 10.1038/s41577-023-00853-1. Epub 2023 Mar 31. PMID: 37002288; PMCID: PMC10064963.

32. Hammarlund E, Lewis MW, Hansen SG, Strelow LI, Nelson JA, Sexton GJ, Hanifin JM, Slifka MK. Duration of antiviral immunity after smallpox vaccination. Nat Med. 2003 Sep;9(9):1131–7. doi: 10.1038/nm917. Epub 2003 Aug 17. PMID: 12925846.

33. Mayr A, Hochstein-Mintzel V, Stickl H, Abstammung, Eigenschaften und Verwendung des attenuierten Vaccinia-Stammes MVA. Infection. 1975 3, 6–14. 10.1007/BF01641272

34. Moodie Z, Price L, Gouttefangeas C, Mander A, Janetzki S, Löwer M, Welters MJ, Ottensmeier C, van der Burg SH, Britten CM. Response definition criteria for ELISPOT assays revisited. Cancer Immunol Immunother. 2010 Oct;59(10):1489–501. doi: 10.1007/s00262-010-0875-4. Epub 2010 Jun 15. PMID: 20549207; PMCID: PMC2909425.

35. Butler A, Hoffman P, Smibert P, Papalexi E, Satija R. Integrating single-cell transcriptomic data across different conditions, technologies, and species. Nat Biotechnol. 2018 Jun;36(5):411–420. doi: 10.1038/nbt.4096. Epub 2018 Apr 2. PMID: 29608179; PMCID: PMC6700744.

36. Gerritsen B, Pandit A, Andeweg AC, de Boer RJ. RTCR: a pipeline for complete and accurate recovery of T cell repertoires from high throughput sequencing data. Bioinformatics. 2016 Oct 15;32(20):3098–3106. doi: 10.1093/bioinformatics/btw339. Epub 2016 Jun 20. PMID: 27324198; PMCID: PMC5048062.

