## Supplementary Figures for "Viral-based individualized neoantigen vaccine as adjuvant treatment in resected head and neck squamous cell carcinoma: immunogenicity and efficacy from a randomized Phase I trial"

**Supplementary Table 1. Definition of analysis sets**

| Population | Definition per protocol | Primary dataset | Total number of patients (A/B) |
|---|---|---|---|
| **Full analysis set** (FAS) | All randomized patients | Demographic and baseline characteristics | **N=33** (17/16) |
| **Safety population** (SAF) | All randomized patients who received **at least one injection** of TG4050 | Safety analysis | **N=19** (17/2) |
| **Per protocol** (PP) | Minimum exposure before recurrence: Arm A: patients who **completed their 6 weekly injections** of TG4050 Arm B: patients who **completed their 6 weeks FU** | DFS | **N=32** (16/16) |

**Supplementary Table 2. Next Generation Sequencing Tumor Profiling (Tumor Portrait™)**

| Arm | Patient ID | Treated | PDL1 | TMB | TME functional class | Active Immune infiltration | Tumor cell proliferation |
|---|---|---|---|---|---|---|---|
| Arm A | 02-0101005 | Yes | 38 | 2.3 | Fibrotic | MODERATE | Medium |
| Arm A | 02-0101009 | Yes | 23 | 1.37 | Immune enriched, non-fibrotic | HIGH | Medium |
| Arm A | 02-0101010 | Yes | 29 | 1.99 | Immune enriched, fibrotic | MODERATE | Low |
| Arm A | 02-0102002 | Yes | 28 | 3.19 | Immune Desert | MODERATE | Medium |
| Arm A | 02-0102011 | Yes | <1 | 1.99 | Immune Desert | MODERATE | Low |
| Arm A | 02-0102024 | Yes | 24 | 3.42 | Immune Desert | MODERATE | Medium |
| Arm A | 02-0102025 | Yes | <1 | 4.2 | Immune Desert | LOW | Medium |
| Arm A | 02-0102033 | Yes | 80 | 7.7 | Immune enriched, fibrotic | MODERATE | Medium |
| Arm A | 02-0102039 | Yes | 48 | 1.46 | Immune Desert | MODERATE | Low |
| Arm A | 02-0102040 | Yes | 43 | 1.68 | Immune enriched, non-fibrotic | MODERATE | Medium |
| Arm A | 02-0602001 | Yes | 81 | 4.34 | Immune enriched, non-fibrotic | MODERATE | Medium |
| Arm A | 02-0602004 | Yes | 27 | 3.28 | Immune Desert | LOW | Medium |
| Arm A | 02-0602008 | Yes | 24 | 1.9 | Immune enriched, non-fibrotic | MODERATE | Low |
| Arm A | 02-0602009 | Yes | 19 | 3.16 | Fibrotic | MODERATE | Medium |
| Arm A | 02-0602013 | Yes | 26 | 4 | Immune enriched, non-fibrotic | MODERATE | Low |
| Arm A | 02-0602016 | Yes | <1 | 3.05 | Immune Desert | LOW | Medium |
| Arm A | 02-0602021 | Yes | 48 | 2.41 | Immune Desert | LOW | High |
| Arm B | 02-0101001 | No | 0 | 2.1 | Immune Desert | LOW | Medium |
| Arm B | 02-0101006 | Yes | 44 | 2.91 | Immune enriched, non-fibrotic | MODERATE | Low |
| Arm B | 02-0101008 | No | 90 | 3.28 | Immune enriched, non-fibrotic | HIGH | High |
| Arm B | 02-0101011 | No | <1 | 1.9 | Immune Desert | MODERATE | Medium |
| Arm B | 02-0101013 | No | <1 | 0.03 | Immune enriched, non-fibrotic | MODERATE | Medium |
| Arm B | 02-0102007 | Yes | <1 | 3.56 | Immune Desert | MODERATE | Low |
| Arm B | 02-0102018 | No | <1 | 4.26 | Immune Desert | LOW | Medium |
| Arm B | 02-0102019 | No | <1 | 1.6 | Immune Desert | MODERATE | Medium |
| Arm B | 02-0102020 | No | 15 | 3.02 | Immune Desert | MODERATE | Medium |
| Arm B | 02-0102027 | No | 17 | 3.64 | Immune Desert | LOW | Medium |
| Arm B | 02-0102030 | No | <1 | 2.77 | Immune Desert | MODERATE | Medium |
| Arm B | 02-0602005 | No | <1 | 3.02 | Immune Desert | MODERATE | Medium |
| Arm B | 02-0602010 | No | 1 | 3.36 | Immune Desert | MODERATE | Medium |
| Arm B | 02-0602011 | No | 72 | 5.24 | Immune Desert | MODERATE | Medium |
| Arm B | 02-0602017 | No | 48 | 7.95 | Fibrotic | MODERATE | Medium |
| Arm B | 02-0602018 | No | 50 | 0.34 | Immune Desert | MODERATE | Low |

| | | | Neoantigen | | | | MVA | |
|---|---|---|---|---|---|---|---|---|
| Arm | Patient_ID | Vaccine neoantigens | ELISpot | Tetramer | Overall neoantigen responses | Overall response | ELISpot | Tetramer |
| A | 02-0101005 | 26 | Not tested | 1/23 | 1 | De novo | Not tested | De novo |
| A | 02-0101009 | 30 | Not tested | 1/11 | 1 | De novo | Not tested | Not tested |
| A | 02-0101010 | 30 | 0/30 | 0/20 | 0 | No response | Amplified | Amplified |
| A | 02-0102002 | 27 | 3/27 | 1/12 | 4 | De novo | No response | Not tested |
| A | 02-0102011 | 30 | 1/30 | 1/9 | 1 | De novo | Amplified | Not tested |
| A | 02-0102024 | 30 | Not tested | Not tested | N/A | N/A | Not tested | Not tested |
| A | 02-0102025 | 30 | 1/30 | 2/12 | 3 | De novo | De novo | Not tested |
| A | 02-0102033 | 30 | 1/30 | 2/12 | 3 | De novo | De novo | De novo |
| A | 02-0102039 | 30 | 1/30 | 0/5 | 1 | Amplified | De novo | Not tested |
| A | 02-0102040 | 17 | 0/17 | 0/4 | 0 | No response | De novo | Not tested |
| A | 02-0602001 | 30 | 2/18 | 2/19 | 3 | De novo | De novo | Not tested |
| A | 02-0602004 | 30 | 0/30 | 1/18 | 1 | De novo | De novo | Not tested |
| A | 02-0602008 | 30 | 0/30 | 0/16 | 0 | No response | De novo | Not tested |
| A | 02-0602009 | 30 | Not tested | Not tested | N/A | N/A | Not tested | Not tested |
| A | 02-0602013 | 30 | 7/30 | 0/29 | 7 | De novo | Amplified | Not tested |
| A | 02-0602016 | 30 | 0/30 | Not tested | 0 | No response | Stable | Not tested |
| A | 02-0602021 | 30 | 16/30 | 5/18 | 16 | De novo | De novo | De novo |
| | Total Arm A | | 8 / 13 (61.5%) | 9 / 14 (64.3%) | 11 / 15 (73.3%) | | | |
| | Median in treatment responders | | 2 [1-16] | 1 [1-5] | 3 [1-16] | | | |
| B | 02-0101006 | 30 | Not tested | 0/10 | 0 | | Not tested | Not tested |
| B | 02-0102007 | 30 | Not tested | 2/33 | 2 | | Not tested | Not tested |

**Supplementary Figure 1. Overview of T cell responses between baseline and D64**

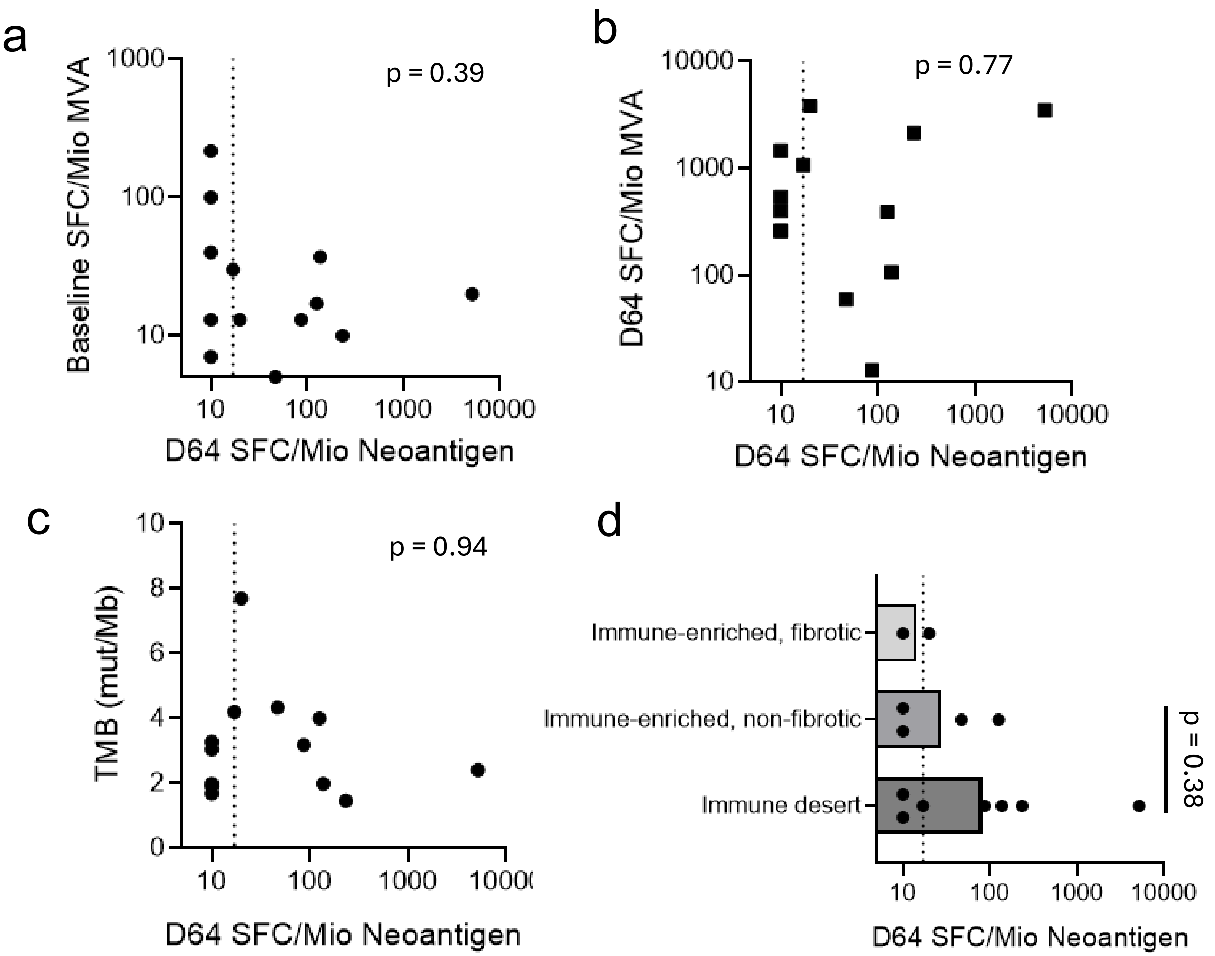


**Supplementary Figure 2. Anti-MVA T cell responses at baseline or D64, TMB or TME subtype are not associated with the levels of neoantigen-specific T cell responses after treatment.** Neoantigen-specific T cell responses, calculated as the sum of background-subtracted spot-forming cells (SFC) per million PBMC across all responding neoantigens at day 64 (D64) and measured by *ex vivo* IFN-γ ELISpot, were plotted against:(a) background-subtracted SFC per million PBMC in response to the vaccinia/MVA peptide pool at baseline,(b) background-subtracted SFC per million PBMC in response to the vaccinia/MVA peptide pool at D64,(c) tumor mutational burden (TMB, mutations per Mb), and(d) tumor microenvironment (TME) subtype.

Statistical analysis: p-values were obtained using non-parametric tests on log-transformed values. Spearman's rank correlation was applied for panels (a–c). For panel (d), a Mann–Whitney test was performed between TME categories with ≥3 samples per group.

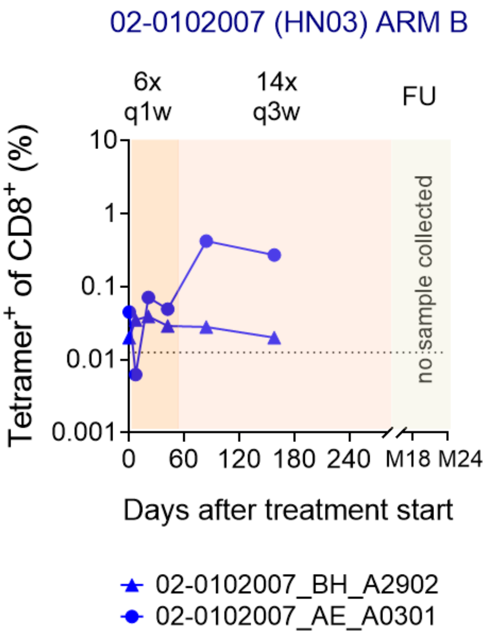


**Supplementary Figure 3. Neoantigen-specific CD8+ T cell responses in one Arm B patient treated at recurrence**

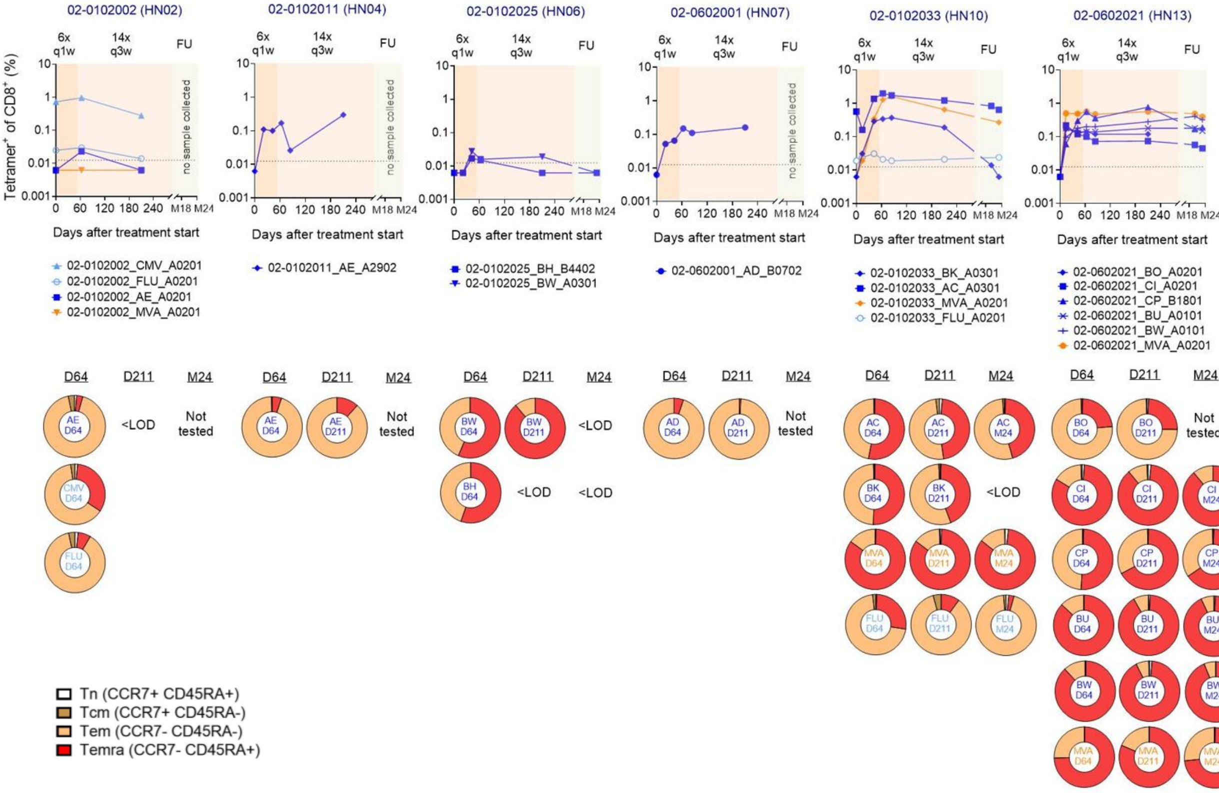


**Supplementary Figure 4. Phenotype of neoantigen-specific and virus-specific CD8⁺ T cells detected by tetramer in Arm A patients**

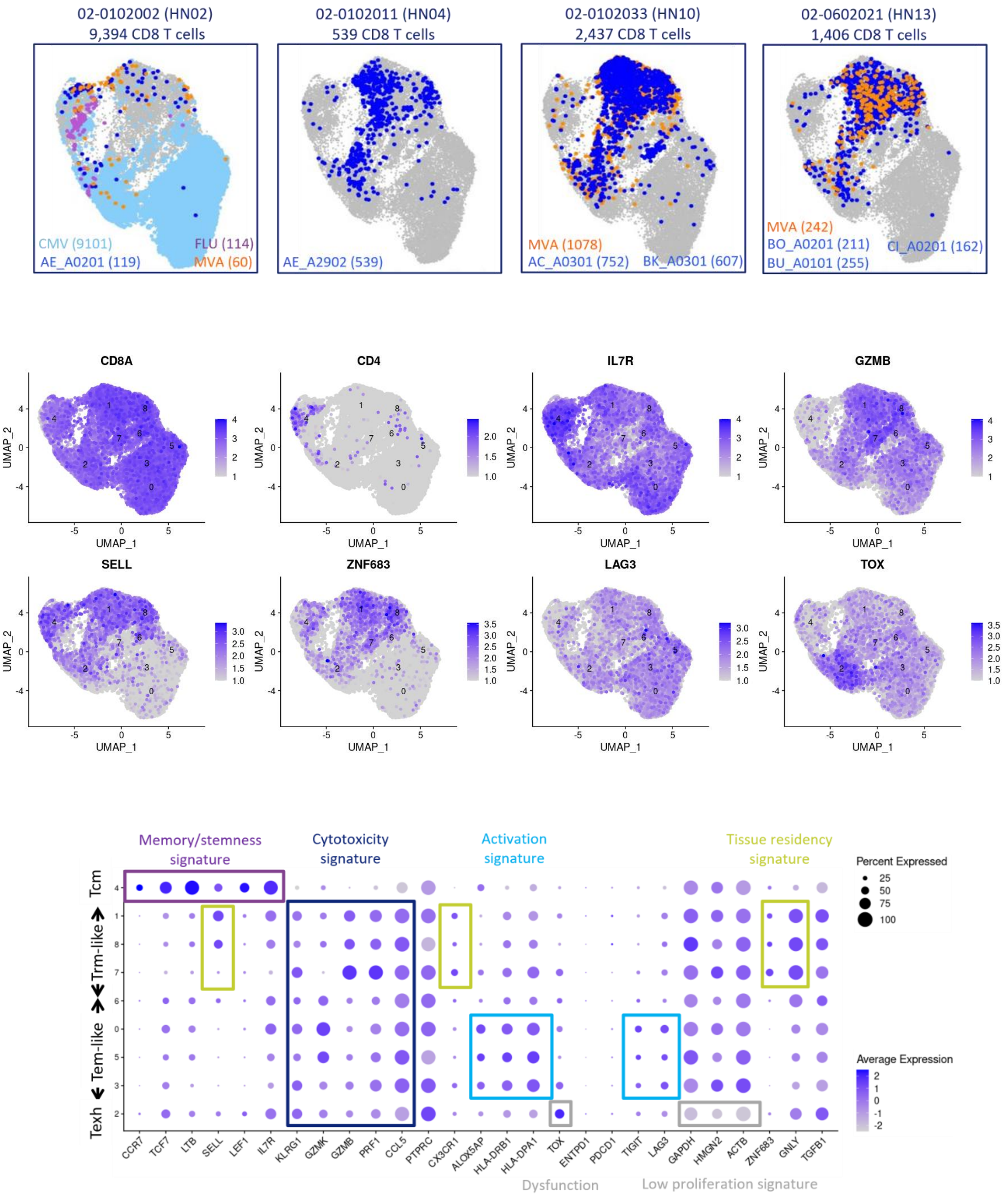


**Supplementary Figure 5. scRNAseq clusters**

**Supplementary Figure 6. Expansion of tumor-infiltrating T cell clones after TG4050 treatment**

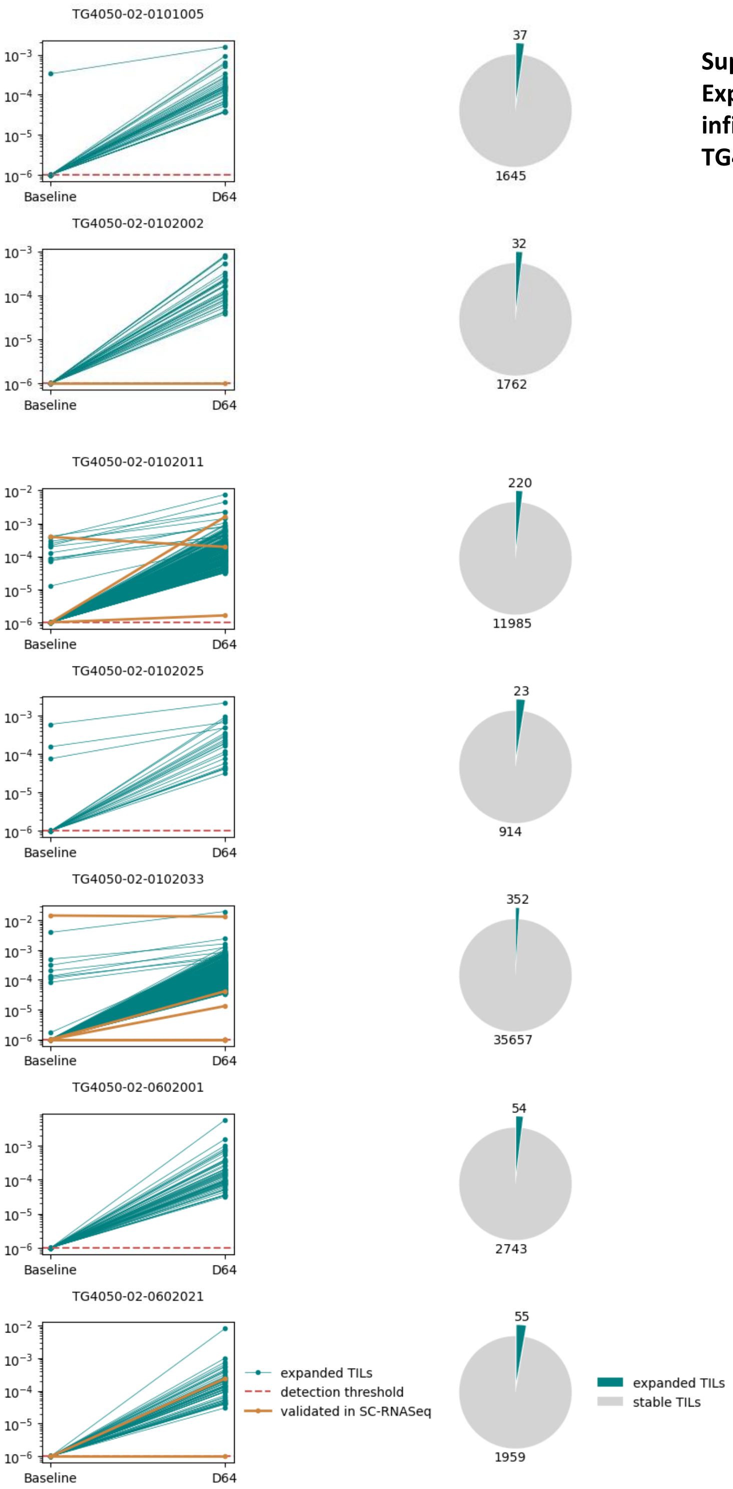

**Supplementary Figure 7 . Schematic representation of TG4050 expression cassettes**

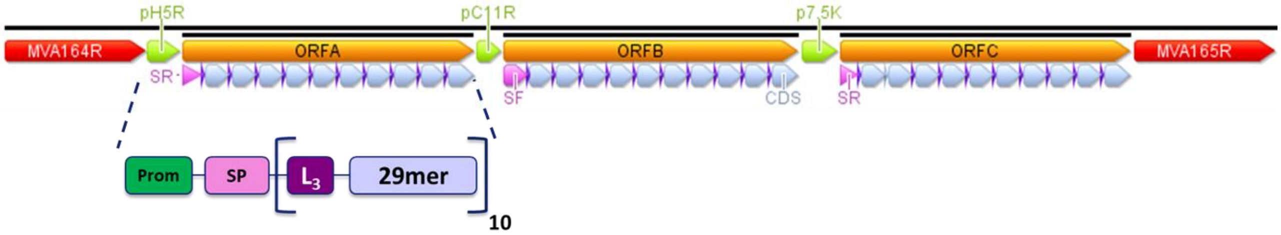


Prom : promoter (pH5R, pC11R, p7.5K)

SP : Signal peptide of Rabies glycoprotein (SR) or of Measles F glycoprotein (SF)

$L_3$ : linker: GSG, GAS, GTS

29mer : Neoepitope (29aa)

MVA : Region homologous to MVA164R and 165R genes
